# Spatial mapping of the hepatocellular carcinoma landscape identifies unique intratumoural perivascular-immune neighbourhoods

**DOI:** 10.1101/2023.09.15.23295451

**Authors:** Felix Marsh-Wakefield, Cositha Santhakumar, Angela L. Ferguson, Thomas M. Ashhurst, Joo-Shik Shin, Fiona H. X. Guan, Nicholas J. Shields, Barry J. Platt, Givanna H. Putri, Ruta Gupta, Michael Crawford, Carlo Pulitano, Charbel Sandroussi, Jerome M. Laurence, Ken Liu, Geoffrey W. McCaughan, Umaimainthan Palendira

## Abstract

**Background & Aims:** Hepatocellular carcinoma (HCC) develops in the context of chronic inflammation, however, the opposing roles the immune system plays in both the development and control of tumours is not fully understood. Mapping immune cell interactions across the distinct tissue regions could provide greater insight into the role individual immune populations have within tumours.

**Methods:** A 39-parameter imaging mass cytometry panel was optimised with markers targeting immune cells, stromal cells, endothelial cells, hepatocytes, and tumour cells. We mapped the immune landscape of tumour, invasive margin, and adjacent non-tumour regions across sixteen resected tumours comprising of 144 regions of interest. X-shift clustering and manual gating were used to characterise cell subsets, and Spectre quantified the spatial environment to identify cellular neighbourhoods. Ligand-receptor communication was quantified on two single-cell RNA-sequencing datasets and one spatial transcriptomic dataset.

**Results:** We show immune cell densities remain largely consistent across these three regions, except for subsets of monocyte-derived macrophages which are enriched within the tumours. Mapping cellular interactions across these regions in an unbiased manner identifies immune neighbourhoods comprised of tissue-resident T cells, dendritic cells, and various macrophage populations around perivascular spaces. Importantly, we identify multiple immune cells within these neighbourhoods interacting with VEGFA^+^ perivascular macrophages. *VEGFA* was further identified as a ligand for communication between perivascular macrophages and CD34^+^ endothelial cells.

**Conclusions:** Immune cell neighbourhood interactions, but not cell densities, differ between intratumoural and adjacent non-tumour regions in HCC. Unique intratumoural immune neighbourhoods around the perivascular space points to an altered landscape within tumours. Enrichment of VEGFA^+^ perivascular macrophages within these tumours could play a key role in angiogenesis and vascular permeability.

**Impact and Implications:** We investigated the landscape of immune cells within liver cancer. A unique perivascular neighbourhood of immune cells was identified. This is important as the interaction of immune cells and the vasculature in liver cancer is a therapeutic target of current systemic therapy. The characterisation in detail of this neighbourhood may potentially identify additional targets for future therapies.

**Highlights:**

- Imaging mass cytometry using 39 markers was used to map immune-infiltrating cells in hepatocellular carcinoma (HCC)
- Perivascular macrophages (PVM) were seen to form unique intratumoural neighbourhoods with immune cells
- PVM were identified as VEGFA^+^ within HCC tumours

## Introduction

Liver cancer is the sixth most incident cancer and the third leading cause of cancer-related death in the world ^1^ with hepatocellular carcinoma (HCC) accounting for approximately 90 % of cases ^2^. The five-year overall survival of HCC after diagnosis is approximately 20 % and this statistic has not changed significantly over the past decade despite advancements in treatment options ^3^. This poor prognosis is attributable to its phenotypic and genetic heterogeneity, its occurrence in patients with reduced liver reserve (cirrhosis), high rates of tumour recurrence following curative treatments, and its predisposition to metastasise in advanced stages ^4^. Although immunotherapy has recently revolutionised the treatment of advanced-stage disease in other malignancies such as melanoma and lung cancer, objective response rates to systemic combination therapies in advanced HCC have been suboptimal (less than 30 %) ^5–7^.

Being the prototypical cancer of inflammatory origin, immune responses in HCC are implicated in both pro- and anti-tumoural activities ^8^. Understanding the complex interplay between immune cells and the tumour environment is therefore critical. Recent studies using single-cell approaches have identified the diverse phenotype of immune cells within HCC tumours and provided some insight into their possible functional roles ^9^. However, several of these single-cell methodologies lack spatial context, therefore limiting our ability to determine the potential impact that specific cellular interactions have on clinical outcomes. Spatially resolving these cellular interactions could provide greater insight into how various immune populations contribute to tumour development, progression, and control.

Angiogenesis is also a key feature of HCC, wherein hypervascularisation and leaky vessels pose challenges to the cellular immune control of tumours and influences responses to various therapies ^10–12^. Amongst many angiogenic factors, vascular endothelial growth factor A (VEGFA) has been identified as the key driver of neovascularisation in HCC, with tumour cells largely implicated in its production ^13,14^. The importance of angiogenesis (and its interplay with the tumour immune microenvironment, TME) is evident by recent data showing that combination bevacizumab (anti-VEGFA) and atezolizumab (anti-PD-L1) antibodies improved survival in patients with advanced unresectable HCC ^5^. Interestingly, this strategy has been found to have a synergistic effect ^15,16^.

HCC tumour cells are a well-known source of VEGFA ^13,17^, yet several immune cells have been identified as alternate sources of VEGFA. *VEGFA*^+^ mast cells are thought to be pro-tumour across several cancer types, although they are largely absent in HCC ^18^. In contrast, high levels of *FOLR2*^+^ macrophages, *PLVAP*^+^ endothelial cells, and Notch/VEGF signalling are associated with an immunosuppressive niche within HCC tumours ^19^.

High-dimensional imaging mass cytometry (IMC) has previously been utilised to investigate the tumour microenvironment in HCC across several contexts, including hepatitis B virus infection ^20^, response to immunotherapies ^21^, and macrophage characterisation ^22^. The latter study is important as several studies have implicated macrophages in HCC, including neighbourhood interactions associated with immune cell activation ^23,24^ and response to immunotherapies ^21^. However, more work is needed to better understand the role of immune cells – particularly macrophages – in angiogenesis and vascular permeability in HCC.

In this study, we interrogated the tumour and adjacent non-tumour immune microenvironment in patients with resectable HCC using a high-dimensional tissue imaging platform known as IMC. We spatially resolved cellular interactions in these regions to identify immune neighbourhoods in intratumoural perivascular spaces that comprised of liver-resident CD4^+^ and CD8^+^ T cells, dendritic cells, and phenotypically diverse macrophages. Importantly, we showed enrichment of VEGFA^+^ perivascular macrophages (PVMs) within the tumour regions that interact with these immune aggregates in the perivascular space that was absent in adjacent non-tumour regions, suggesting these immune neighbourhoods could be important in the development and/or progression of HCC.

## Methods

### Patients

Institutional human research ethics was obtained by the Sydney Local Health District Ethics Review Committee (HREC 2020/ETH02093). Research was conducted in accordance with both the Declarations of Helsinki and Istanbul, with written consent given in writing by all subjects. Treatment naïve patients undergoing curative resection for HCC from 2019 until 2021 at a single tertiary institution were identified from the Royal Prince Alfred Liver Biobank. Inclusion criteria was based on the availability of liver tissue (frozen or paraffin embedded) from adult (>18 years old) males and females with HCC diagnosis. Patients also required availability of clinical, imaging, and histopathology data. The formalin-fixed paraffin embedded (FFPE) tissues were retrieved from the Department of Tissue Pathology and Diagnostic Oncology and Royal Prince Alfred Hospital. Relevant clinicopathological data was obtained from electronic medical records. Clinicopathological data included age, sex, aetiology of liver disease, presence of cirrhosis, presence of microvascular invasion (MVI), and largest tumour size. Of the sixteen HCC patients, two have since begun treatment with immune checkpoint inhibitors, and one is now part of a placebo-controlled trial. Whole specimens of HCC tissues from an additional eight patients were used to assess the intratumoural spatial relationship of PVMs and endothelial cells. Further details are provided in Supplementary Table 1.

### Tissue Microarray Preparation

The haematoxylin and eosin (H&E) stained sections from the sixteen patients were reviewed by a liver pathologist. One region of interest (ROI) was selected from the tumour, invasive margin, and non-tumour regions. The tumour ROI was selected based on the high abundance of tumour-infiltrating lymphocytes (TILs) on H&E stained sections. The invasive margin was defined at the border of the malignant tumour and consisted of approximately 50 % malignant and 50 % non-malignant tissue. The non-tumour region was selected away from the tumour. The exact distance between the non-tumour and tumour was variable across patient samples with no fixed diameter. In many patient samples, these areas were selected from different blocks so we are unable to attain a fixed measurement for this distance. Three 1 mm diameter (triplicate) cores were taken from each ROI for creation of the two tissue microarrays (TMAs) (Figure 1A). Tonsils and hepatocellular adenoma were used as batch controls for the TMAs. Both TMAs were sectioned at 7 μm onto charged slides. Figure 1 provides an overview of the experimental design.

**Figure 1.**
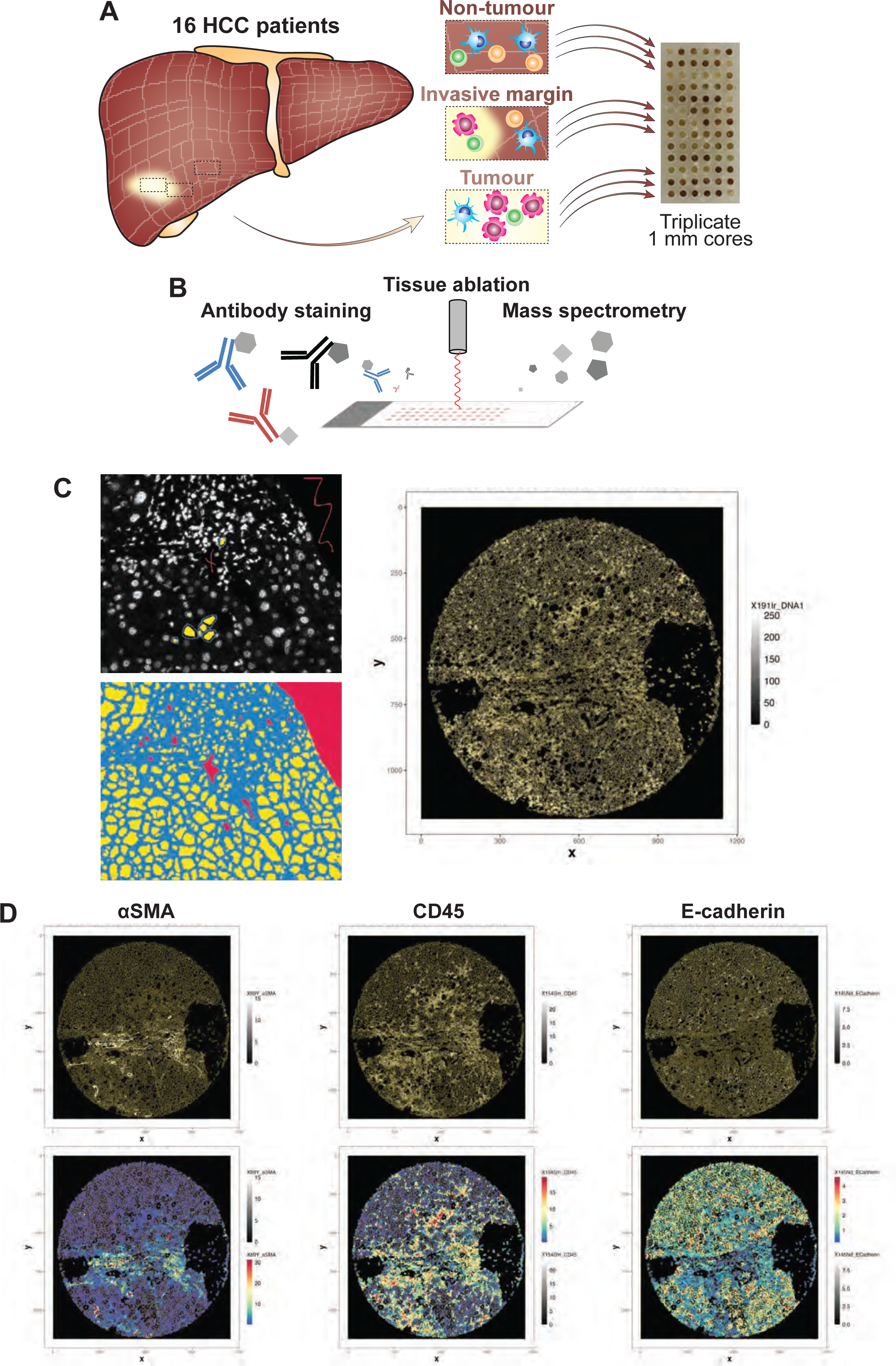
Experimental overview. A) Sixteen HCC patients underwent liver resections. 1 mm diameter regions of non-tumour, invasive margin, and tumour were selected from each patient in triplicate to create a tissue microarray. B) Sections were stained with antibodies conjugated to heavy metals which were then ablated by a Helios imaging mass cytometer to quantify regions. C) Ilastik software was used to define cell borders to create a mask of single cells. D) R package Spectre quantified marker expression for each segmented cell across regions.

### Antibody Panel

An antibody panel (Supplementary Table 2) was designed to identify various components of the HCC TME and background liver tissue, including malignant and non-malignant hepatocytes, immune cells, immune checkpoint inhibitors, cell signalling pathways, stromal cells, and structural markers. Antibodies were selected based on clones previously validated in our lab on human tissue ^25^. Antibodies were then re-validated on FFPE liver and HCC tissue (Supplementary Figure 1). Antibodies that were purchased unconjugated were all tested and validated by conventional immunohistochemistry (IHC) prior to metal conjugation. Post-metal conjugation antibodies were revalidated and titrated using IMC. Pre-conjugated antibodies (Sydney Cytometry Reagent Bank) were tested and titrated for our tissue and staining conditions prior to experimental use. Antibody-metal conjugations were done using the X8 Maxpar conjugation kit as per the manufacturer’s protocol (Standard BioTools).

### Tissue staining for imaging mass cytometry

TMA sections were baked at 60 °C for 60 minutes prior to deparaffinisation and rehydration. Heat-induced antigen retrieval was performed by boiling the samples at 100 °C for 15 minutes in pH 9.0 antigen retrieval buffer (10 mmol L^−1^ Tris base, 1 mmol L^−1^ EDTA, 0.05 % Tween 20, pH 9.0). The TMA slides were allowed to cool down to room temperature (RT) before proceeding.

Sections were incubated with 3 % hydrogen peroxide prior to washing and blocking (Akoya Biosciences, Antibody Diluent/Block). Slides were then stained with anti-CD69 (clone EPR21814, Abcam) at RT before washing with TBS-T. Secondary OPAL-HRP (PerkinElmer) was added to the slides at RT before washing in TBS-T. Slides were stained with TSA Plus FITC (PerkinElmer) at RT before washing with TBS-T. Slides were microwave treated (as described above) before letting them cool down to RT before proceeding.

Slides were then washed in TBS-T before blocking in blocking buffer (Akoya Biosciences) for 45 minutes at 37 °C. Slides were blocked with 20 % donkey serum (in DPBS) for 10 minutes at RT. Slides were stained with anti-PD-L1 (clone E1L3N, CST) and anti-CK8/18 (clone 5D3, Leica) for 45 minutes at RT before washing in TBS-T. Slides were then stained with Cy3 anti-rabbit (711-166-152, Jackson ImmunoResearch Laboratories) for PD-L1 and Cy5 anti-mouse (715-606-150, Jackson ImmunoResearch Laboratories) for CK8/18. Slides were washed in TBS-T and DPBS. Slides were avidin/biotin blocked (Life Technologies). Slides were then stained in anti-Foxp3 biotin (clone 236A/E7, eBioscience) before washing in TBS-T. TMA sections were incubated overnight with a metal-tagged antibody cocktail (Supplementary Table 2, including anti-FITC, anti-Cy3, anti-Cy5, and anti-biotin) at 4 °C.

TMA sections were washed in 0.1 % Triton-X (in DPBS) and DPBS. TMA sections were stained with Cell-ID Ir-Intercalator in DPBS (Standard BioTools). TMA sections were washed in deionised H_2_O before being allowed to air dry at RT (Figure 1B).

### Imaging Mass Cytometry

The Hyperion imaging mass cytometer (Standard BioTools) was used to acquire data from both TMA slides. A pulsed laser scanned and laser ablated the tissue at 200 Hz. IMC data files were analysed using MCD Viewer (version 1.0.560.6, Standard BioTools). MCD Viewer and histoCAT++ (version 2.2)^26^ were used for visualising the images, TIFF extraction, and creating representative pseudo-colour images. (Figure 1C-D).

### Multiplex Immunohistochemistry

FFPE tumour specimen blocks were sectioned at 4 μm and stained using OPAL multiplex IHC (mIHC) staining kit (Akoya Biosciences) according to optimised in-house protocols as previously described ^27,28^. Tissue sections were baked at 60 °C for 60 minutes prior to deparaffinisation and rehydration. Heat-induced antigen retrieval was performed by boiling the samples at 100 °C for 15 minutes in pH 9.0 antigen retrieval buffer. Sections were incubated with 3 % hydrogen peroxide prior to washing and blocking (Antibody Diluent/Block, Akoya Biosciences). Sections were then incubated with a single purified primary antibody for 35 minutes at RT, washed, and then incubated with HRP (OPAL polymer HRP (Akoya biosciences), sheep-HRP (Invitrogen), or MACH-3 mouse 2-step (Biocare, 10 minutes/step)) for 10 minutes at RT. Sections were washed prior to incubation with OPAL fluorochromes diluted in tyramide signalling amplification (TSA) reagent (Akoya Biosciences). Antigen retrieval was repeated, and subsequent antibodies stained for as described above. At completion of all antibody staining, samples were counterstained for DAPI (Cell Signalling Technologies) prior to mounting with Prolong Diamond (Life Technologies).

Primary antibodies used were anti-HLA-DR (TAL-1B5, Abcam), anti-FXIIIA (SAF13A, Affinity Biologicals), anti-CD34 (QBEnd/10, Leica), and anti-VEGFA (VG1, Dako). Single marker staining is shown in Supplementary Figure 2.

Images (20X for quantification and 20X or 40X for representative images) from 8 HCC specimens within tumour regions were captured using the Mantra quantitative pathology imaging system in combination with Mantra Snap (version 1.0.3, Akoya Biosciences) and inForm (version 2.4.2, Akoya Biosciences) to spectrally process images. Multispectral images were exported as TIFF single images and were then imported into Fiji (version 1.53c) ^29^ to process representative images. HALO (version 3.6.4134, Indica Labs) was used to perform cell segmentation, threshold markers for cell phenotyping, and spatial analysis. Proximity analysis was performed within the HALO Spatial Analysis module (Indica Labs). Phenotyped cells were registered and plotted together on a single plot. The Proximity Analysis tool was used to identify the number of VEGFA^+/–^ macrophages within a proximity range of 0-100 μm of CD34^+^ endothelial cells and non-endothelial cells in 20 μm increments.

### Spatial data Analysis

Ilastik software (version 1.4.0b27) was used to segment single cells using the multicut method ^30^, as this method does not rely on nuclei (Figure 1C). Masks were created to then quantify marker expression levels for each single cell in R (version 4.2.0)^31^ using ‘Spectre’ (version 1.0 and development version)^32^ (Figure 1D).

Single cells were manually gated using FlowJo (version 10.8, BD Bioscience) to identify conventional immune cell subsets (Supplementary Figure 3). The area of each section was calculated using Fiji (version 1.53c) ^29^, which was then used to calculate the density of each conventional subset. Spectre was then used to quantify, for every cell, the average number of each cell subset within a 20 µm radius.

For unsupervised analysis, X-shift clustering (version <u>26-Apr-2018</u>) was used ^33^, which identified 238 clusters (Supplementary Figure 4A). The median signal intensity of CD45 was calculated for each cluster before running z-score normalisation. Clusters with values below zero were considered CD45^−^, and values above zero were considered CD45^+^. The latter were divided into CD45^low^ and CD45^hi^ with a cut-off equal to the absolute value of the smallest value (Supplementary Figure 4B). Cells then underwent X-shift clustering within each of the three groups (CD45^−^, CD45^low^, and CD45^hi^) resulting in 91, 36, and 18 clusters respectively. Similar clusters were then combined using the function simprof as part of the ‘clustsig’ R package (version 1.1)^34^, generating 47 clusters (20 CD45^+^ clusters and 27 CD45^−^ clusters) (Supplementary Figure 4C). The spatial data were then quantified using Spectre ^32^, including cluster densities, average distance between each cluster and every other cluster, average number of cells within 20 µm for each cluster, and the proportions of each cluster. As each region was done in triplicate, the results were averaged so each patient tissue region had a single value for each quantification.

When analysing the CD45^+^ clusters, the number of clusters with ≥ 5 cells in each region were calculated. Clusters with < 5 cells across less than a third of total regions were excluded, which removed six clusters (Supplementary Figure 4D).

Heatmaps were generated using Spectre. The median signal intensity for each cluster was calculated and then min-max scaled.

Each patient region contained thousands of parameters, so linear dimensionality reduction algorithms were used to reduce the complexity. A principal component analysis (PCA) was performed using Spectre to identify differences between the three tissue ROIs: non-tumour, invasive margin, and tumour. A PCA reduced the dimensions based on the overall variance within a given dataset, including human and experimental variability. In addition to a PCA, a sparse partial least squares-discriminant analysis (sPLS-DA) was done ^35^. An sPLS-DA reduces the dimensions based on differences between groups and is not as strongly affected by unwanted variability. The R package ‘mixOmics’ was used for the sPLS-DA (version 6.24.0)^36^. The sPLS-DA used leave-one-out validation, calculating the Mahalanobis distance with at least three components generated. When using all clusters, up to ten variables were selected for each component. For select immune cell neighbourhood interactions, there was no limit to the number of variables assessed for each component.

### Ligand-receptor communication

Single-cell transcriptomic analyses were done using the GSE149614 dataset (https://www.ncbi.nlm.nih.gov/geo/query/acc.cgi?acc=GSE149614), which consisted of cells from paired non-tumour and tumour (8/10 patients) from newly diagnosed HCC patients ^37^. These patients were pathologically confirmed and proven to have no other cancers. 7/10 patients had viral infection (five with HBV, two with HCV), 3/10 had tumour-node-metastasis (TNM) stage I, 1/10 had stage II, 2/10 stage IIIA, 2/10 stage IIIB, and 2/10 stage IV ^37^. Pre- processed and pre-annotated data were used. Ligand-receptor communication between cells was calculated using the R package ‘CellCall’ (version 1.0.7)^38^.

A second cohort of HCC patients were similarly analysed as a validation (https://data.mendeley.com/datasets/skrx2fz79n/1), which similarly consisted of cells from paired non-tumour and tumour regions from six HCC patients ^39^. 6/6 had HBV infection. Cluster annotations from the first cohort were transferred to this second cohort using ‘scANVI’ as part of ‘scvi-tools’ (version 1.0.4) ^40,41^ (Supplementary Figure 5). Ligand-receptor communication was then similarly calculated on the transferred clusters.

### Spatial transcriptomic analysis

Further confirmation was undertaken using publicly available CosMx SMI data made available by nanoString ^42^. The data contained one HCC tumour (grade G3, stage II) and one normal liver tissue. Data had undergone cell segmentation and cell annotation. Data were analysed in R using vignette from nanoString ^42^.

Endothelial cells were defined based on pre-annotated data, consisting of central venous liver sinusoidal endothelial cells (LSEC), periportal LSEC, and portal endothelial cells. Macrophages consisted of non-inflammatory and inflammatory macrophages. *CD34*-expressing endothelial cells and *VEGFA*-expressing macrophages were identified based on transcript expression. Close cells were defined as being within 20 µm of each other. Ligand-receptor communication was quantified as described above.

### Statistical tests

Statistics were calculated using packages available within R ^31^, using Type III Sum of Squares, unless stated otherwise. Plots were generated using R and GraphPad Prism (version 9.0.0). For the PCA, a permutational multivariate analysis of variance (PERMANOVA) was done. The data were scaled with the Euclidean distance calculated between datapoints. Permutational tests are powerful non-parametric tests that do not assume homogeneity of variance or normality of distribution ^43^. 4,999 permutations (for a total of 5,000 tests) were done using all parameters to compare between groups (non-tumour, invasive margin, and tumour regions). The R package ‘vegan’ was used for PERMANOVA calculations (version 2.6-4)^44^. To calculate paired p-values between groups, the R package ‘pairwiseAdonis’ was used with Holm’s correction for multiple comparisons (version 0.4.1)^45^. A similar method was used for the sPLS-DA, but only the parameters that were identified in the first two components were used in their calculation.

For the manually gated results, a Friedman test with Dunn’s multiple comparison corrections was done to compared between groups (non-tumour, invasive margin, and tumour regions). For comparisons between non-tumour and tumour, a Wilcoxon test was used. These were calculated in Graphpad Prism (version 9.0.0).

For comparisons between non-tumour and tumour, a PERMANOVA with Holm’s correction was used for the sPLS-DA (as above). For paired comparisons for individual parameters a permutation student’s t-test was done using the perm.t.test function as part of the ‘RVAideMemoire’ R package (version 0.9-83)^46^.

### Results Study design

Sixteen treatment naïve HCC patients who underwent liver resections were included in this study. Of these patients, twelve were male and the median age was 64.5 years (range 33-84 years). The aetiology of chronic liver disease was viral in seven patients, non-viral in eight patients, and dual viral and non-viral pathology in one patient. Nine patients were cirrhotic, and half the cases had evidence of microvascular invasion. The median largest tumour size was 36 mm (range 9-270 mm). Quantitative protein expression on single cells and spatial mapping of cellular neighbourhood interactions were performed on patient tissue samples from selected regions (non-tumour, invasive margin, and tumour) (Figure 1C-D).

### The spatial landscape differs between HCC tumour regions

We mapped the full cellular landscape (immune cells, stromal cells, endothelial cells, hepatocytes, and tumour cells) of HCC to identify any differences between non-tumour, invasive margin, and tumour regions. X-shift clustering was initially performed on all cells. This unbiased approach identified 47 clusters (Figure 2A). The spatial environment was quantified for each patient and tissue region. A PCA was run on > 10,000 parameters, including individual cluster levels (density, percentage of total cells, count), average distances between clusters, and neighbour interactions across each region. Cellular neighbourhoods were inferred based on the average number of one cluster within 20 µm of another (or same) cluster ^47^. The PERMANOVA revealed significant differences between the three regions (p=0.0030), which was limited to a difference between non-tumour and tumour (p=0.0006) but not between invasive margin and tumour or non-tumour (Figure 2B, p=0.2467). We then determined the parameters that contribute most to the identified variance. This revealed the primary differentiator as the distance between individual clusters, suggesting that the spatial distribution of cells could be key in HCC (Figure 2C). Using a more targeted approach that provides greater granularity to the parameters that potentially differentiate these regions, the sPLS-DA also showed clear differences between each of the three regions (Figure 2D). The differentiating parameters included not only distance between clusters, but also cell proportions and densities (Figure 2E). Component 1 (represented by the x-axis) separated all regions. Cluster 37, identified as endothelial cells, had differences in density, percentage, and count. Cluster 34, another endothelial cell subset, differed in percentage and count. Cellular neighbourhood interactions of cluster 44 (myeloid cells) to 16 (hepatocytes), cluster 12 (hepatocytes) to 34 (endothelial cells), and cluster 12 (hepatocytes) to 37 (endothelial cells) also differed between regions, particularly between non-tumour and tumour regions. Component 2 (y-axis) separated the invasive margin, with cellular neighbourhood interactions of cluster 3 (T cells) to 28 (endothelial cells). Together, these data show that our unbiased analytical approach to identify differences in HCC tissue regions demonstrates clear distinctions in the overall cellular landscape. We were particularly interested in the immune cell microenvironment within these regions. Thus, we specifically examined quantities and spatial relationships of immune cells.

**Figure 2.**
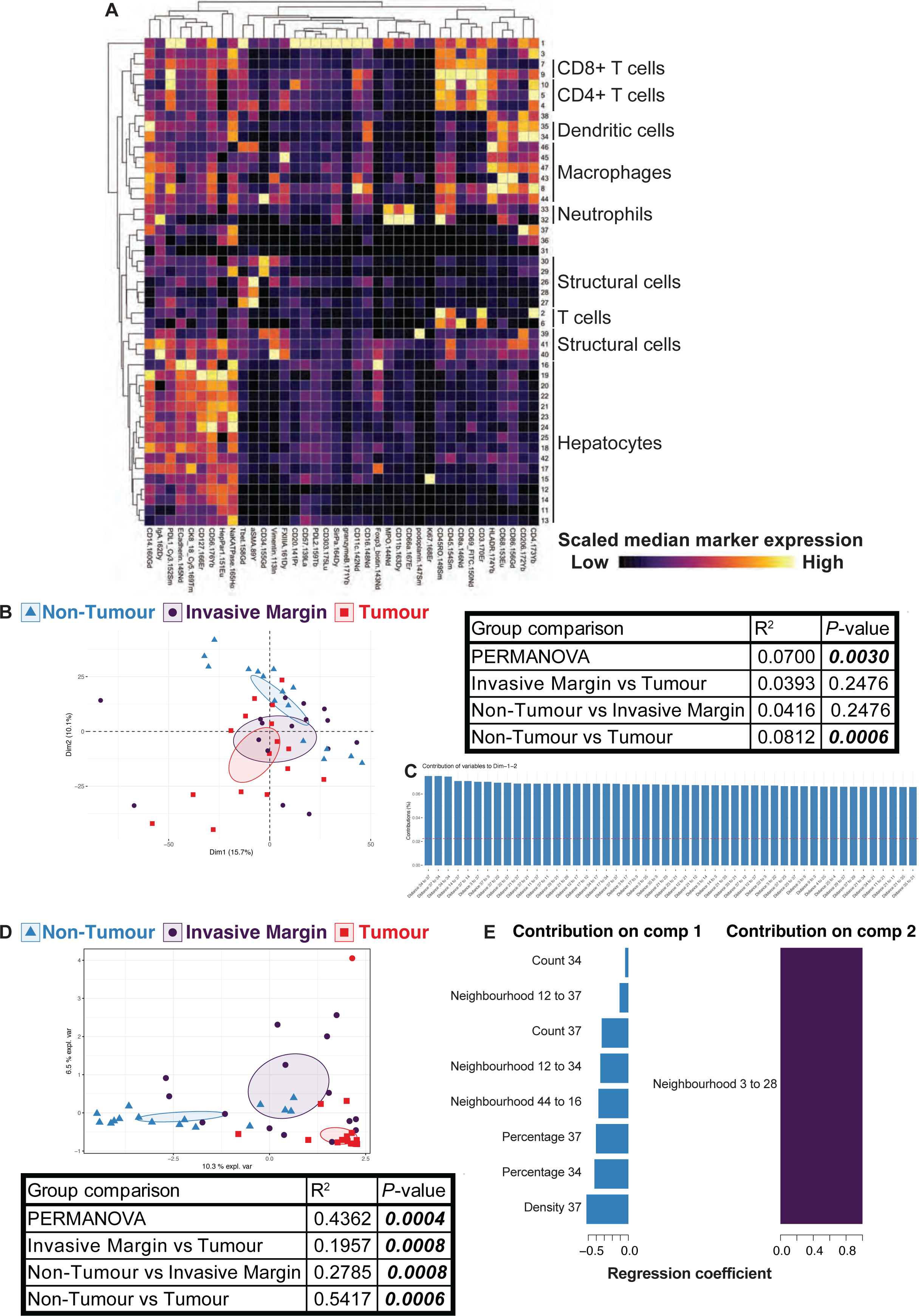
Spatial environment differs between liver regions. A) X-shift clustering was used to identify 47 clusters represented by a heatmap. Each row is a different cluster, each column a different marker. B) Principal component analysis (PCA) of all spatial data. Symbols and colours represent a different region from individual patients. 95 % confidence interval shown. A PERMANOVA with Holm’s correction was performed on all parameters. C) The parameters that contributed most to the variance across the first two components are shown. Horizontal red dashed line represents the value if all parameters contributed equally. D) A sparse partial least squares-discriminant analysis (sPLS-DA) generated a similar plot, with the selected parameters for the first two components shown. A PERMANOVA with Holm’s correction was performed on select parameters from the first two components. E) The parameters that contribute most to the differences between regions across the first two components are shown. n = 16 HCC patients, each with non-tumour, invasive margin, and tumour regions.

### Innate, but not adaptive, immune cell densities differ between HCC and adjacent tissue

Differences in the immune landscape of the tumour, invasive margin, and adjacent non-tumour regions were investigated to determine whether cell densities differentiated the three regions. Manual gating analysis was performed to identify conventional immune cell subsets, including both innate and adaptive immune populations (Figure 3). Surprisingly, the densities of these immune subsets were mostly similar across the three regions. T cells (particularly CD4^+^ T cells) constituted the largest proportion of immune cells across all regions and their densities remained consistent (Figure 3A-B, Supplementary Figure 6A). Of the myeloid cells, macrophages constituted the largest proportion across regions (Figure 3A). There was a higher density of macrophages and dendritic cells within tumour compared to non-tumour regions (Figure 3B).

**Figure 3.**
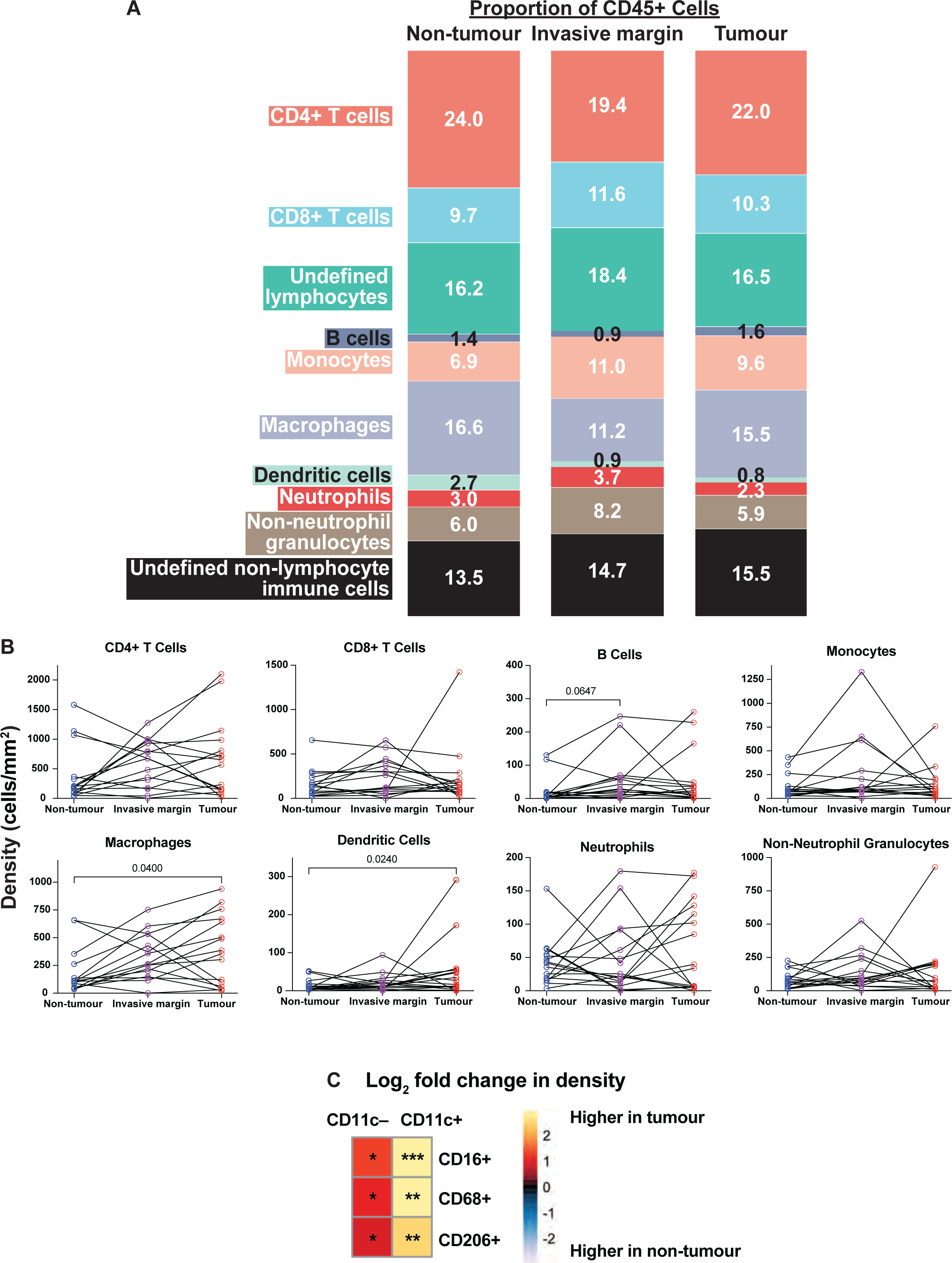
Tumour has higher densities of monocyte-derived CD11c^+^ and CD11c^−^ macrophage subsets compared to non-tumour regions. A) Proportion of conventional immune cells across regions. B) The density of conventional immune subsets across regions. Friedman test with Dunn’s multiple comparison corrections was performed. C) Log_2_ fold change comparing densities of monocyte-derived CD11c^+^ and CD11c^−^ macrophages between tumour and non-tumour regions. Wilcoxon test was used. *p<0.05, **p<0.01, ***p<0.001. n = 16 HCC patients.

A more targeted analysis of innate immune cell subsets revealed subsets of macrophages (CD16^+^ CD68^+^ CD206^+^ CD11c^+/–^), possibly monocyte-derived (based on CD14 expression), were significantly higher within tumour compared with non-tumour regions (Figure 3C). In contrast, CD11c^+^ and CD11c^−^ macrophages that were CD14^−^ were not different (Supplementary Figure 6B). These subsets were also higher (as a proportion of total cells) in the tumour compared with non-tumour regions, with the exception of monocyte-derived CD206^+^ CD11c^−^ macrophages (Supplementary Figure 6C). As a proportion of total immune cells, only the monocyte-derived CD11c^+^ macrophages were higher in the tumour compared with non-tumour regions (Supplementary Figure 6C).

There were no significant differences in the density of T cell subsets between regions (Figure 3B). Most T cells were CD4^+^ rather than CD8^+^ across regions, with most CD4^+^ T cells expressing CD69, consistent with tissue-resident CD4^+^ T cells (Supplementary Figure 6A).

Together, these data demonstrate that innate immune cells, particularly monocyte-derived macrophages, are significantly increased within tumour when compared to non-tumour regions.

### Unique immune cell neighbourhood interactions exist within the HCC tumour

As the densities and proportions of most conventional immune cell subsets were similar across regions, we next spatially analysed these regions to investigate immune cell neighbourhood interactions. When investigating CD45^+^ clusters alone to focus on immune cells, six clusters had < 5 cells in more than one third of the images, so were removed from further analysis (Supplementary Figure 4D). Furthermore, certain clusters were phenotypically similar to each other based on marker expression and were combined (Supplementary Figure 7). As a result, ten CD45^+^ clusters were included in the neighbourhood analysis (Figure 4A).

**Figure 4.**
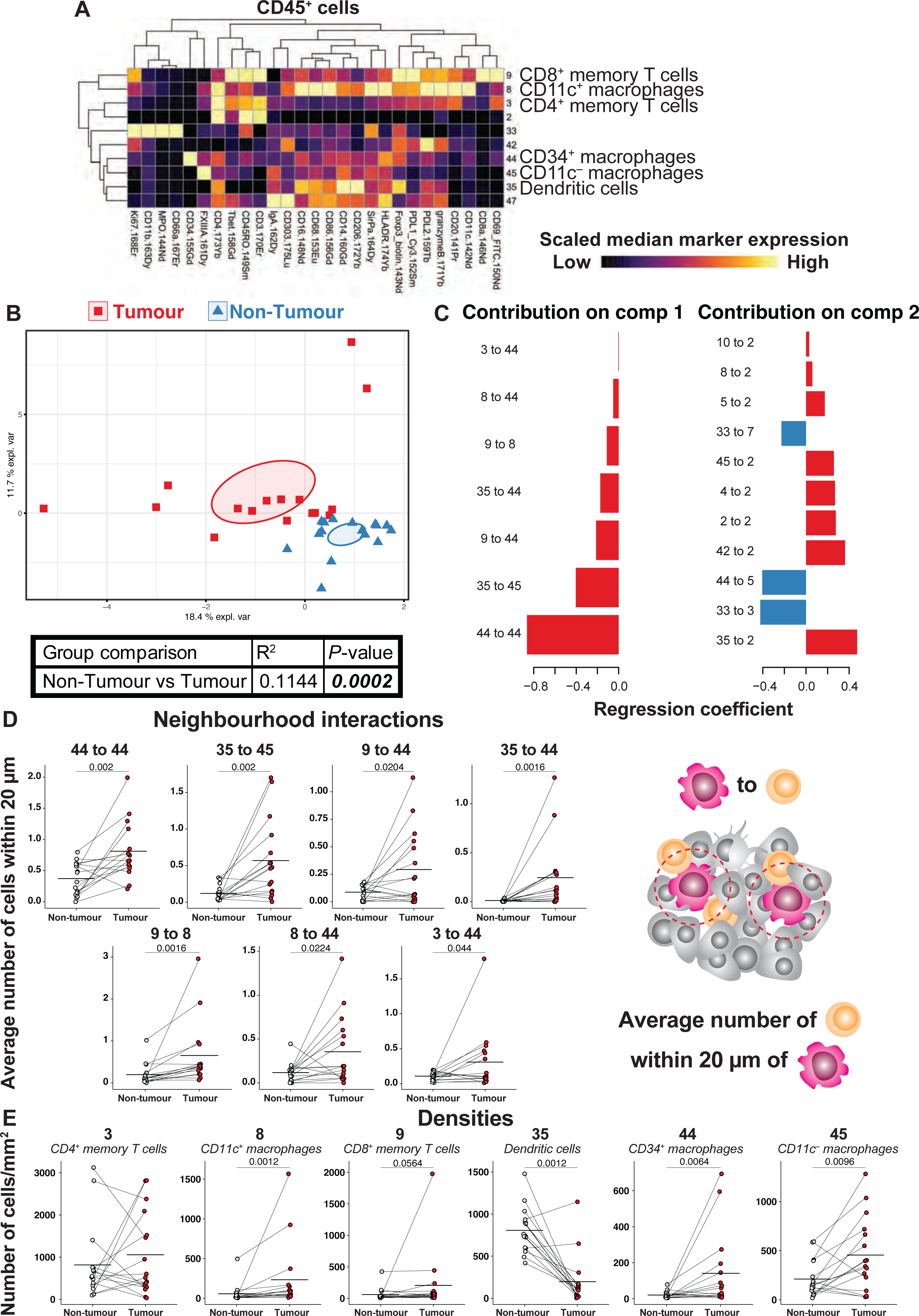
Immune cell neighbours differ between non-tumour and tumour regions. A) Heatmap of CD45^+^ clusters. Each row is a cluster, each column is a marker. B) sPLS-DA comparing non-tumour to tumour regions. Symbols and colours represent a different region from individual patients. 95 % confidence interval shown. C) Selected parameters for the first two components are shown. Select comparisons between non-tumour and tumour cellular neighbourhood interactions (D) and densities (E). n = 16 HCC patients, each with non-tumour, invasive margin, and tumour regions. A permutation student’s t-test was done, p-values ≤0.1 are shown, mean is shown.

To identify immune cell interactions that differ between regions, an sPLS-DA was performed that revealed differences in immune cell neighbourhood interactions between non-tumour and tumour regions (Figure 4B). Multiple immune cell neighbourhood interactions were identified (Figure 4C). The first component (x-axis) parameters were further investigated. Cluster 44 was more commonly within 20 µm of other clusters (including 3, 8, 9, 35, 44) within tumour compared to non-tumour regions. Likewise, cluster 35 was commonly within 20 µm of cluster 45, and cluster 9 within 20 µm of cluster 8 within HCC tumour regions. These seven immune cell neighbourhood interactions were identified to be more common within tumour regions when compared to non-tumour regions (Figure 4D). We then determined whether these interactions could have been impacted by individual cell densities. The density of clusters 8, 9, 44, and 45 was higher within tumour compared to non-tumour whilst the opposite was true for cluster 35 (Figure 4E). There was no difference in the density of cluster 3 when comparing tumour and non-tumour regions. These data show that multiple immune cell neighbourhood interactions are more frequent within HCC tumour regions when compared to non-tumour regions. Some of these increased cellular neighbourhood interactions may be attributed to increased densities of the cells involved.

### Increased immune cell interactions with VEGFA^+^ perivascular macrophages in HCC tumour tissue

Having identified cluster 44 at the centre of multiple cellular neighbourhood interactions, this cell cluster was investigated further. Closer examination of the expression markers associated with cluster 44 revealed expression of both CD45 (indicating a haematopoietic cell) and tumour endothelial marker CD34 (Figure 5A). This prompted us to closely examine all images to determine potential close interactions between two different cell types (Supplementary Figure 8). We found that cluster 44 was in fact two discrete cells in close proximity: a tumour endothelial cell characterised by CD34 expression, and a myeloid cell which on further phenotyping also expressed CD14, CD16, CD45, CD68, CD86, CD206, FXIIIA, and HLA-DR, but not CD11c (Figure 5B). Based on the close proximity of this myeloid cell to the tumour endothelial cell with reference to the phenotypic markers expressed, including FXIIIA that is expressed by macrophages ^48–52^, we hypothesised the myeloid cell to be a PVM. Further staining using mIHC, which allowed for greater image resolution, showed that many of these PVM-like cells also expressed VEGFA, suggesting they are indeed PVMs (Figure 5C) ^53–55^. These cells were also identified in additional specimens of HCC tissues from eight patients. Representative images of mIHC staining of four of these HCC tumours are shown in Figure 6C. This staining also showed VEGFA^−^ macrophages in close association to CD34^+^ endothelial cells indicating that both VEGFA^+^ and VEGFA^−^ macrophages preferentially reside in close proximity to CD34^+^ endothelial cells (Figure 6A). Analysis of spatial interactions of VEGF^+/–^ macrophages to CD34^+^ endothelial cells in HCC tumour tissue from these eight patients identified that VEGFA^+^ and VEGFA^−^ macrophages preferentially reside 0-20 μm from CD34^+^ endothelial cells (Figure 6A). Although there was no significant difference in the number of VEGFA^+^ macrophages compared to VEGF^−^ macrophages in the proximity of CD34^+^ endothelial cells, significantly more VEGFA^+^ and VEGFA^−^ macrophages reside in close proximity (0-20 μm from) to CD34^+^ endothelial cells compared to those residing further away, 40-60, 60-80, 80-100 μm (Figure 6A). In contrast, no significant difference in the number of VEGFA^+^ macrophages residing close to non-endothelial cells (0-20 μm) was found when compared to those residing further away (20-100 <u>μ</u>m, 20 μm increments) (Figure 6B).

**Figure 5.**
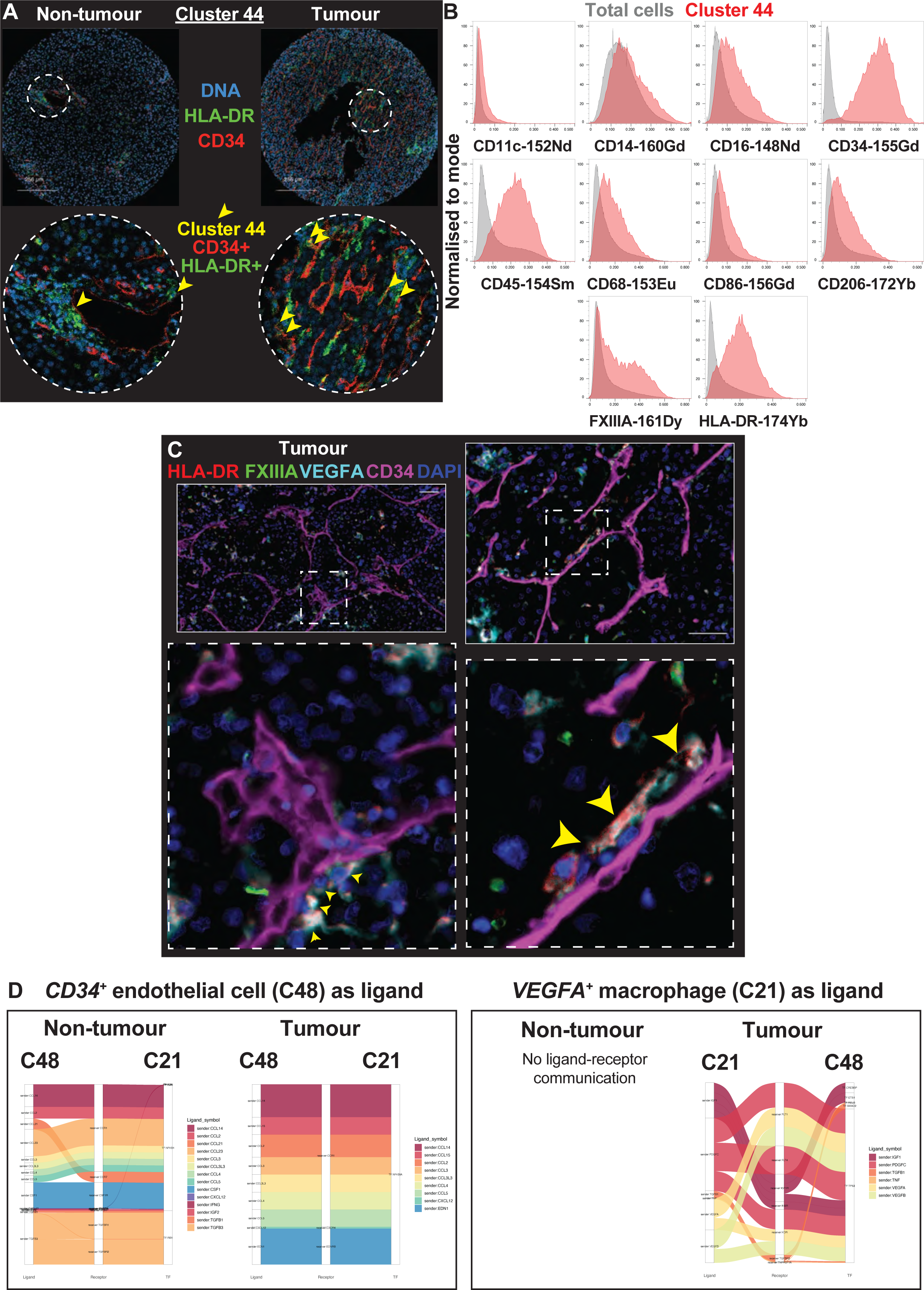
Cluster 44 consists of endothelial and VEGFA^+^ perivascular macrophages. A) Cluster 44 (green) location indicated between non-tumour and tumour regions from the same patient. Yellow arrows point towards the location of HLA-DR^+^ CD34^+^ cells (yellow). B) Histograms showing relative marker expression for cluster 44 (red) compared to total cells (grey). C) OPAL multiplex immunohistochemistry of tumour regions. HLA-DR (red), FXIIIA (green), VEGFA (cyan), CD34 (purple), and DAPI (dark blue). Yellow arrows pointing to white cells expressing HLA-DR, FXIIIA, and VEGFA. D) Ligand-receptor communication between *CD34^+^* endothelial cells and *VEGFA^+^* macrophages. 10 patients with HCC and paired non-tumour and tumour regions were used.

**Figure 6.**
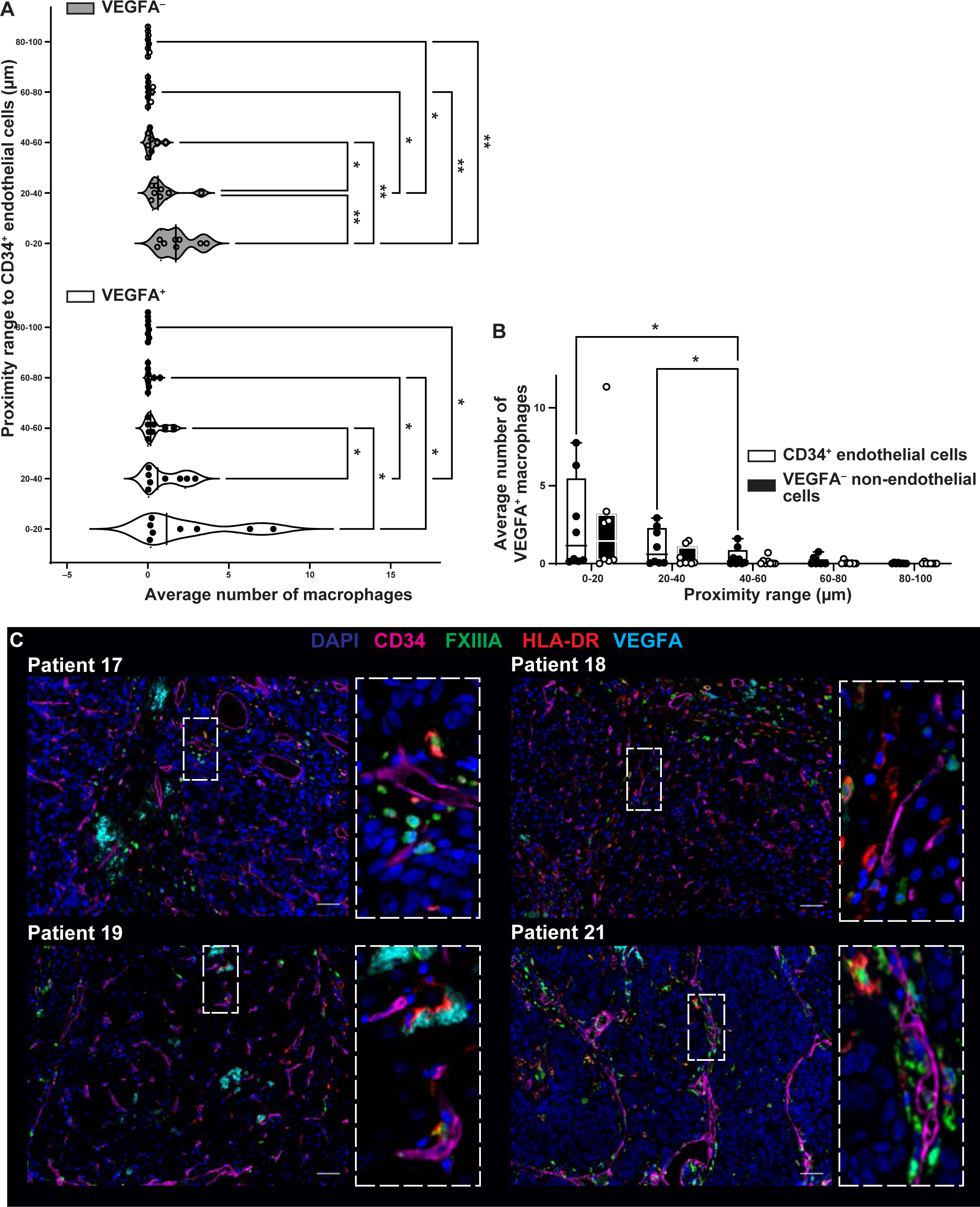
VEGFA^+^ and VEGFA^−^ perivascular macrophages preferentially reside in close proximity to CD34^+^ endothelial cells intratumourally in HCC. A) Waterfall plot of number of VEGFA^+^ and VEGFA^−^ macrophages proximity to CD34^+^ endothelial cells, 20 μm increments, range 0-100 μm. B) Number of VEGFA^+^ macrophages in proximity to CD34^+^ endothelial cells compared to non-endothelial cells (CD34^−^ VEGFA^−^), 20 μm increments, range 100 μm. C) 4/8 HCC patients representative images of OPAL mIHC of HCC tumour tissue stained for HLA-DR (red), FXIIIA (green), VEGFA (cyan), CD34 (magenta), and DAPI (blue). Scale bar= 50 μm. 2-way ANOVA with Uncorrected Fisher’s test was used, *p<0.05, **p<0.001.

**Figure 7.**
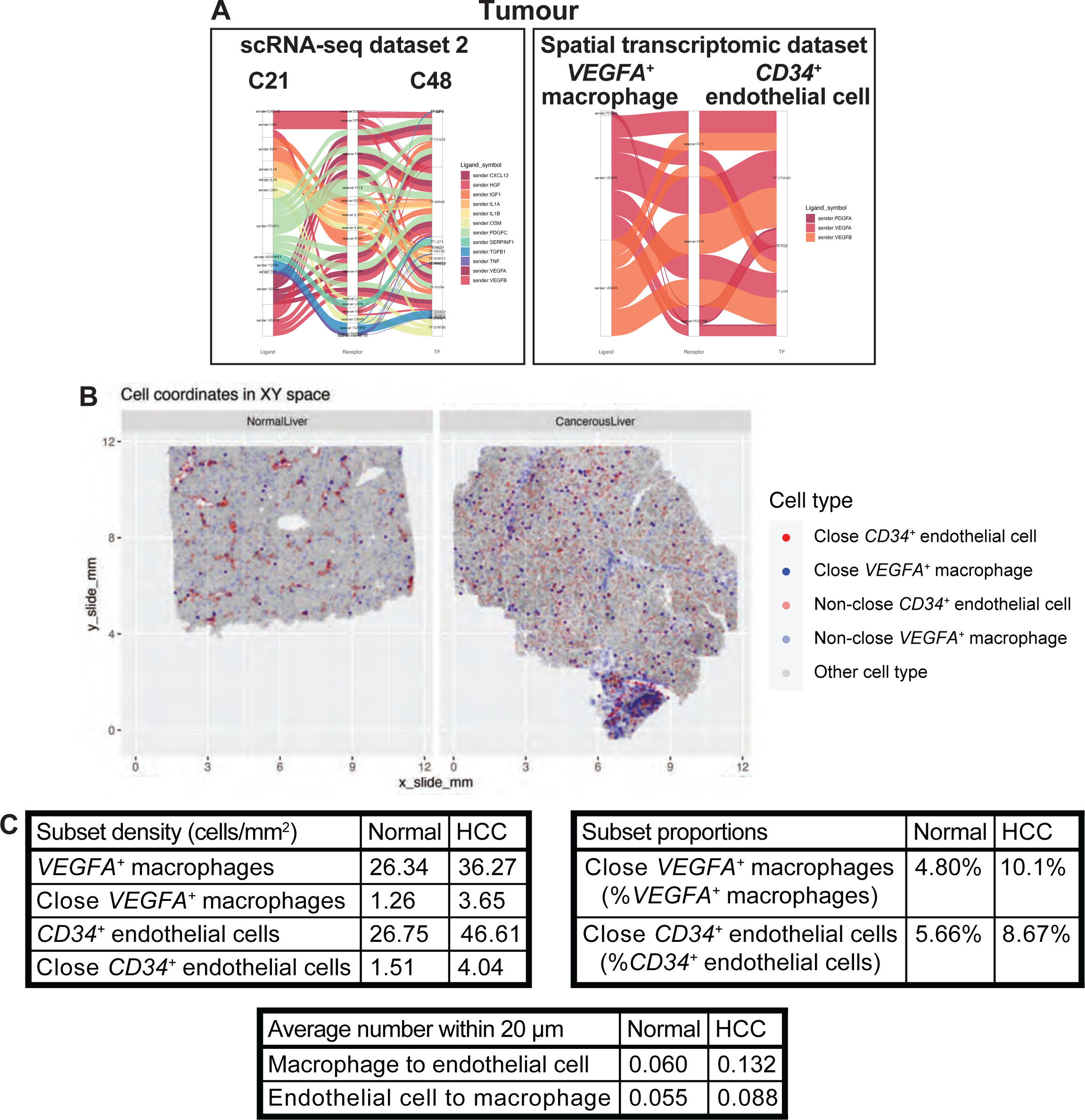
*VEGFA^+^*macrophages and *CD34^+^* endothelial cells have a ligand-receptor communication via *VEGFA* within tumours. A) Ligand-receptor communication between *VEGFA^+^*macrophages (C48, ligand) and *CD34^+^* endothelial cells (C21, receptor) within tumour. The single-cell RNA-sequencing (scRNA-seq) dataset consisted of paired non-tumour and tumour regions from six HCC patients. The spatial transcriptomic dataset consisted of one liver section from one HCC patient. B) Visualisation of the spatial transcriptomic data for normal and HCC liver. *CD34^+^* endothelial cells are red, *VEGFA^+^* macrophages are blue. Close cells (within 20 µm) are solid, whilst non-close cells are translucent. Other cell types are grey. C) Quantification of relative levels of *CD34^+^* endothelial cells and *VEGFA^+^* macrophages in the spatial transcriptomic dataset (*n* = 1).

To determine whether any cell-cell communication was occurring between the CD34^+^ endothelial cells and the VEGFA^+^ PVM, publicly-available single-cell transcriptomic data were analysed. The GSE149614 dataset consisted of tumour resections from newly diagnosed HCC patients with paired non-tumour and tumour tissue cell dissociates ^37^. The pre-annotated data included endothelial cells expressing *CD34* (C48) and macrophages expressing *VEGFA* (C21) that we considered to most closely resemble PVM. Ligand-receptor communication was inferred using ‘CellCall’ ^38^. When C48 (*CD34^+^* endothelial cells) acted as the ligand, several chemokine ligands were predicted to interact with chemokine receptors on C21 in both non-tumour and tumour regions (Figure 5D). When C21 (*VEGFA^+^* macrophages) acted as the ligand, there was communication within the tumour but not non-tumour tissue with endothelial cell (C48) receptors. Furthermore, *VEGFA* and *TGFB1* were predicted ligands from C21, suggesting the VEGFA^+^ macrophages are communicating with CD34^+^ endothelial cells. To expand upon these results, additional validation was done. Similar findings were observed in two additional publicly available cohorts, including a second single-cell RNA-sequencing (scRNA-seq) cohort and a sub-cellular spatial transcriptomic dataset using CosMx (Figure 7A). *VEGFA^+^*macrophages within 20 µm of *CD34^+^*endothelial cells are visualised in normal and cancerous liver (Figure 7B). The levels of *VEGFA^+^* macrophages and *CD34^+^* endothelial cells are summarised in Figure 7C. Despite *n* = 1, there was a higher density and proportion of *VEGFA^+^* macrophages and *CD34^+^* endothelial cells within cancerous liver compared to normal liver, as well as a higher level of cellular neighbourhood interactions (Figure 7C). Together these data reveal that tumour regions are not only highly enriched with CD34^+^ tumour endothelial cells, but also have VEGFA^+^ PVM that potentially interact with these endothelial cells in that region.

### Perivascular immune cell aggregates consist of liver-resident CD4^+^ and CD8^+^ T cells, dendritic cells, and macrophages within HCC tumour tissue

Having identified multiple clusters neighbouring with cluster 44, we then defined all clusters that were found in close proximity of cluster 44. Clusters 3 (Figure 8A) and 9 (Figure 8B) were T cells (CD45^+^ CD3^+^). Further phenotyping showed cluster 3 to be CD4^+^ CD45RO^+^ CD69^+^, suggestive of liver-resident memory CD4^+^ T cells, and cluster 9 to be CD8A^+^ CD45RO^+^ CD69^+^ Tbet^+^ PD-L1^+^, likely liver-resident memory CD8^+^ T cells (Figure 8C). Both clusters had a higher interaction with cluster 44 within the tumour, suggesting an interaction between CD4^+^ and CD8^+^ T cells with PVMs and tumour endothelium.

**Figure 8.**
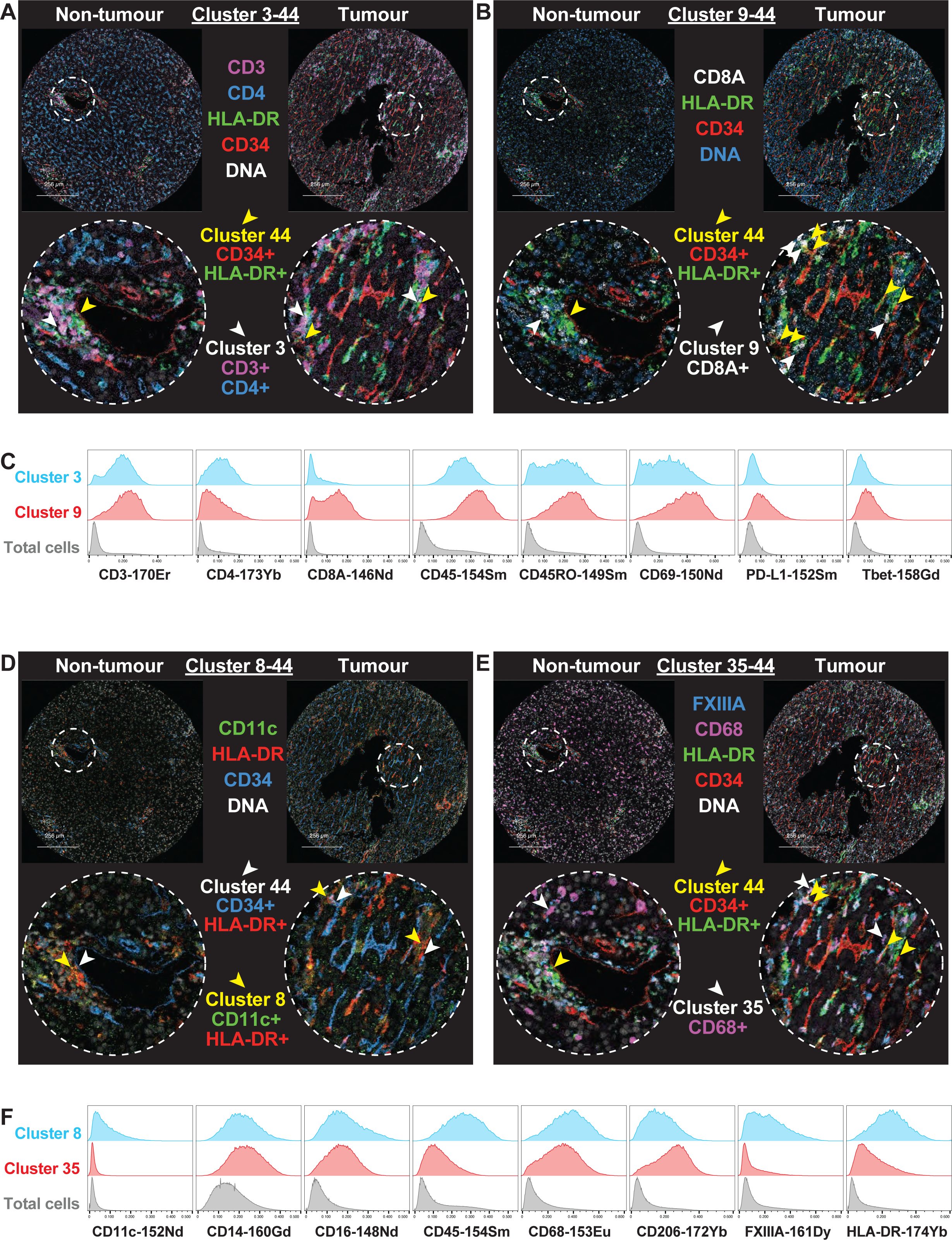
Cluster 44 has a higher number of cellular neighbourhood interactions with clusters 9, 3, 8, and 35 within tumour compared to non-tumour. A) Cluster 3 (CD3^+^ CD4^+^ cells, white arrows) and cluster 44 (CD34^+^ HLA-DR^+^ cells, yellow arrows) location indicated between non-tumour and tumour regions. B) Cluster 9 (CD8A^+^ cells, white arrows) and cluster 44 (CD34^+^ HLA-DR^+^ cells, yellow arrows) location indicated between non-tumour and tumour regions. C) Histograms representing marker expression of cluster 3 (blue), cluster 9 (red), and total cells (grey). D) Cluster 8 (CD11c^+^ HLA-DR^+^, yellow arrows) and cluster 44 (CD34^+^ HLA-DR^+^, white arrows) location indicated between non-tumour and tumour regions. E) Cluster 35 (CD68^+^, white arrows) and cluster 44 (CD34^+^ HLA-DR^+^, yellow arrows) location indicated between non-tumour and tumour regions. F) Histograms representing marker expression of cluster 8 (blue), cluster 35 (red), and total cells (grey). Representative non-tumour and tumour regions are from the same patient.

Clusters 8 (Figure 8D) and 35 (Figure 8E) were CD45^+^ CD14^+^ HLA-DR^+^ myeloid cells. Cluster 8 was also CD11c^+^ CD16^+^ CD68^+^ CD206^+^ FXIIIA^+^, suggestive of a potential macrophage, while cluster 35 was CD11c^−^ CD16^+^ CD68^+^ CD206^+^ FXIIIA^−^, possibly representing a dendritic cell population (Figure 8F). The increased cellular neighbourhood interaction of these cells with PVM-endothelial cell cluster (cluster 44) in the HCC tumour tissue points to an interaction between the tumour endothelium/PVM niche and other myeloid cells within the TME.

Cluster 8, a macrophage subset, was also found to have a cellular neighbourhood interaction with cluster 9, a memory CD8^+^ T cell subset (Figure 9A, Supplementary Figure 9). Clusters 35 (a dendritic cell subset) and 45 were both CD45^+^ CD14^+^ HLA-DR^+^. Cluster 45 had a macrophage phenotype as CD16^+^ CD68^+^ FXIIIA^+^ (Figure 9C). The cellular neighbourhood interactions between these two myeloid cell populations were also higher in the tumour compared to non-tumour regions (Figure 9B) despite the density of cluster 35 being lower in the tumour compared to the non-tumour regions (Figure 4E). Together, these single cell maps reveal that unique intratumoural immune aggregates (consisting of T cells, dendritic cells, and macrophages) interact with PVM and may exert potential pro-tumoural and angiogenic functions. These findings are summarised in Figure 10.

**Figure 9.**
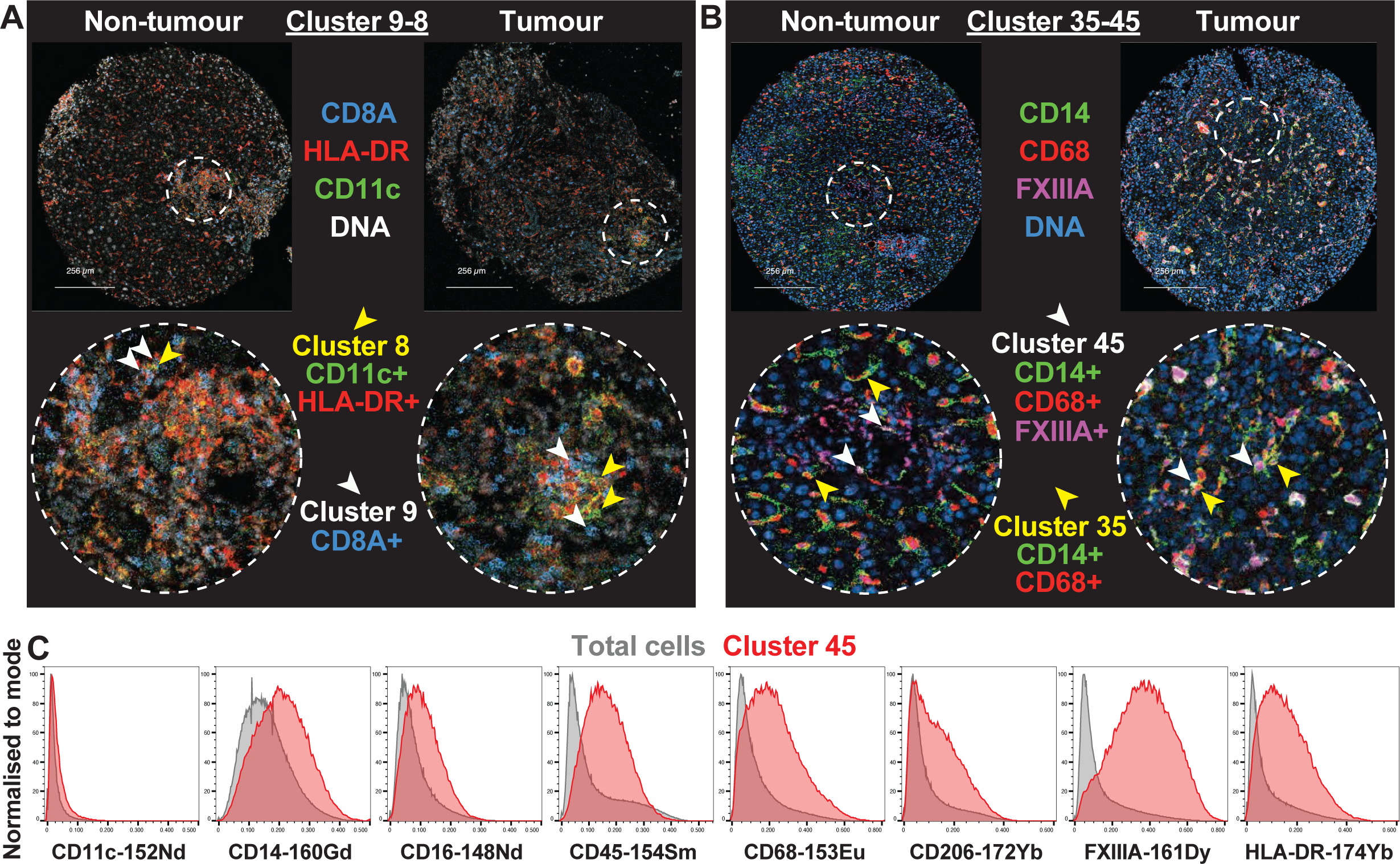
Cellular neighbourhood interactions between clusters 8 and 9, and clusters 35 and 45 are higher in tumour than non-tumour. A) Cluster 8 (CD11c^+^ HLA-DR^+^, yellow arrows) and cluster 9 (CD8A^+^, white arrows) location indicated between non-tumour and tumour regions. B) Cluster 35 (CD14^+^ CD68^+^, yellow arrows) and cluster 45 (CD14^+^ CD68^+^ FXIIIA^+^, white arrows) location indicated between non-tumour and tumour regions. C) Histograms representing marker expression of cluster 45 (red) and total cells (grey). Representative non-tumour and tumour regions are from the same patient.

**Figure 10.**
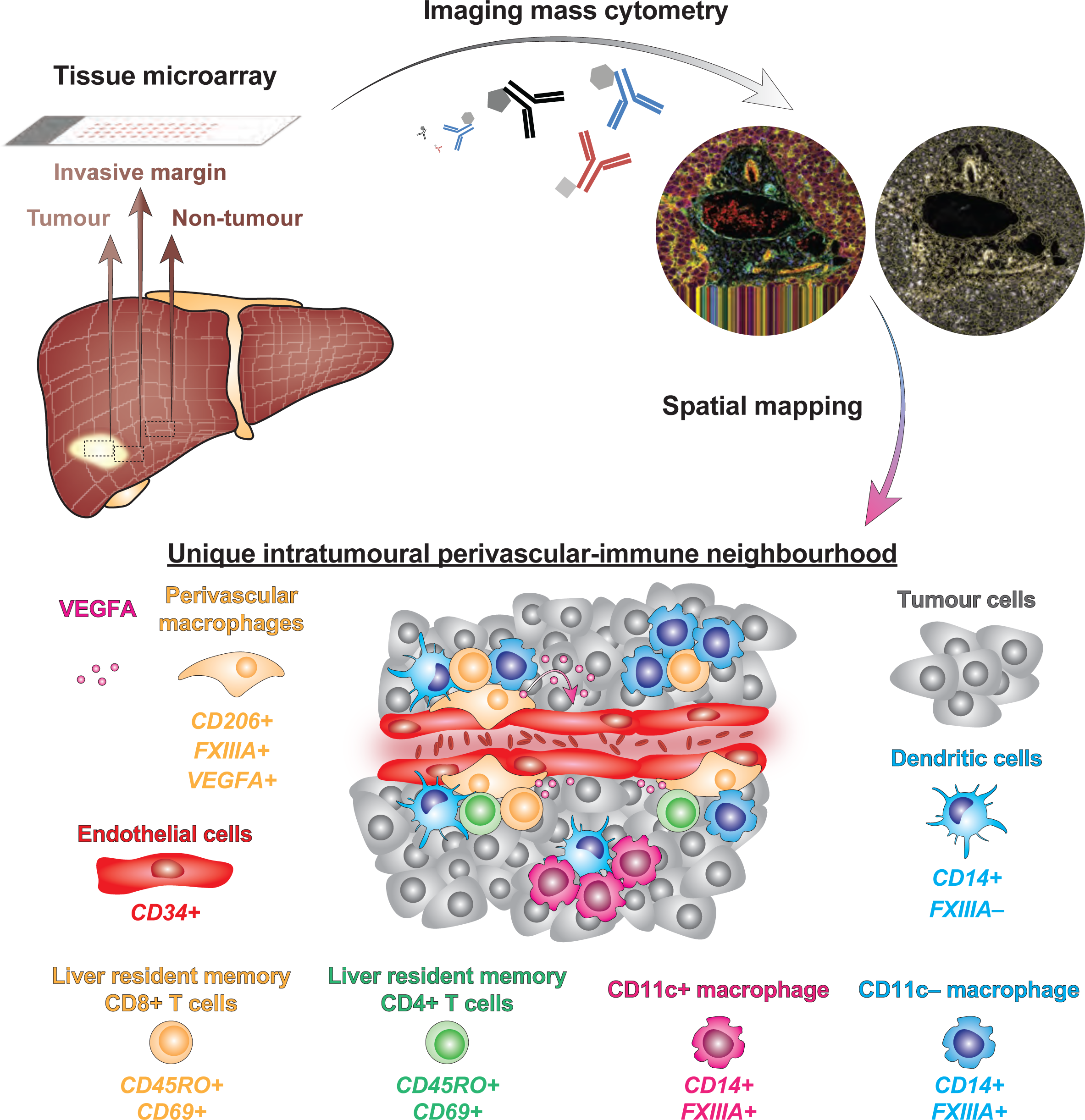
Summary figure. Liver resections were collected from 16 patients with HCC to create a tissue microarray. Regions include non-tumour, invasive margin, and tumour, each collected in triplicate. Imaging mass cytometry was done on sections with a representative image shown. Within the tumour, a unique intratumoural perivascular-immune neighbourhood was identified, featuring cells as described.

Clinical associations were assessed by correlating tumour cellular neighbourhood interactions with prognostic and clinicopathological data. There were no associations between tumour cellular neighbourhood interactions and cirrhosis, early recurrence (<2 years), MVI, or viral-associated HCC (Supplementary Figure 10).

## Discussion

The treatment of HCC remains a major challenge and novel therapeutic strategies are urgently required. Irrespective of its aetiology, HCC mostly develops in diseased liver with concomitant chronic inflammation, thus implicating the immune system in both the pathogenesis of the disease as well as tumour control. Understanding the complex interactions between immune cells and the TME is therefore paramount ^56,57^. We used high-dimensional spatial analysis to map multiple cellular neighbourhood interactions across three regions: the non-tumour, invasive margin, and tumour regions. Mapping the immune landscape across these regions showed that only innate cell densities, but not T cell or B cell densities, are increased within tumour regions compared to non-tumour regions and that macrophages (likely monocyte-derived) are enriched within tumour tissue. We further identified immune aggregates around the perivascular space that are unique to the tumour region. Detailed analysis of this region shows that PVMs are at the centre of these interactions and are closely associated with not only the tumour endothelial cells, but also multiple immune populations, including liver-resident CD4^+^ and CD8^+^ T cells, macrophages, and dendritic cells. Functionally, we show that these PVMs were associated with VEGFA, which is known to be a key driver of angiogenesis and hypervascularity within tumours. Mining publicly available scRNA-seq and spatial transcriptomic datasets to interrogate potential ligand-receptor interactions between PVMs and endothelial cells confirms VEGFA-VEGF receptor interactions and also suggests that other potential pro-tumoural mechanisms involving transforming growth factor-beta (TGF-β) are active within these unique immune aggregates.

How the tumour immune microenvironment and its composition is impacted by hepatocarcinogenesis remains unclear. Our data show enrichment of phenotypically diverse populations of macrophages within the tumour compared to non-tumour regions. Based on CD14 expression, these populations were most likely monocyte-derived ^58,59^. This is in-line with the consensus in the field that tumour-associated macrophages (TAMs) are derived primarily from circulating blood monocytes ^60^, and in human HCC, TAMs arise from CCR2^+^ monocytes ^61^. Interestingly, we found other CD14^−^ macrophage populations were either reduced within tumours or did not show any difference compared to adjacent non-tumour tissue. This, together with no changes in CD8^+^ T cell or CD4^+^ T cell subset densities between tumour and non-tumour regions, suggests that the tumour immune microenvironment may be skewed by the recruitment of monocytes. Furthermore, our findings on several myeloid cell populations regarding their overall numbers and cell-cell interactions should be compared with the findings of Wu *et al.* who devised a myeloid response score (MRS) in using 244 resected human HCC samples ^62^. Tumours with a high MRS score were enriched in CD11b^+^, CD15^+^, CD163^+^, CD33^+^, CD68^+^, and CD204^+^ cells. These were thought to be immunosuppressive in nature and associated with poor prognosis. In addition, Kurebayashi *et al.* found that macrophages were the most common immune cell detected in 919 regions from 158 resected HCC tumours ^63^. CD68 is commonly used as a liver TAM marker ^64^ and this subset was found to be increased in our tumours. Similarly, CD206^+^ CD14^+^ monocyte-derived macrophages were also enriched within the tumour in our study. A dysregulated balance towards CD206^+^ M2 polarised macrophages has also been associated with aggressive tumour phenotypes and worse prognosis in HCC ^65^.

Where these macrophages localise within tumour tissue is critical to our understanding of how the immune environment impacts tumour development, persistence, as well as tumour control. Many previous studies have interrogated the TME in HCC using various techniques including mIHC ^62,63,66^, IMC ^22,67^, and multimodal genomic analysis incorporating scRNA-seq and TCR receptor sequencing ^68–70^. However, not all these techniques facilitate spatial analyses to identify cell-cell interactions between multiple different types of cells within the TME. Our study provides a comprehensive analysis of the composition of the tumour immune microenvironment of HCC and adjacent non-tumour tissue whilst further identifying novel cell-cell interactions between immune cells and the tumour vasculature. Through this spatial mapping we have identified a cellular neighbourhood around the endothelium and PVMs of potential importance.

The close physical association between tumour endothelial cells and macrophages prompted us to define them as PVMs. Currently, there is no consensus on the definition of PVMs as they can have different phenotypes across tissues. However, a unifying feature is that they lie in close proximity to blood vessels, either in direct contact or located within one cell thickness from it ^55^. However, PVMs have not yet been described in healthy adult liver tissue. In diseased tissues such as cancer, these cells have been shown to express VEGFA ^54,55^. We also show that these PVMs are indeed able to produce VEGFA. Importantly, pre-clinical models show that PVMs are also largely derived from circulating monocytes ^54^. In line with this, Matasubra *et al.* found that the majority of angiopoietin (TIE2^+^) expressing macrophages in liver cancer were located in the perivascular areas of tumour tissue and their presence not only correlated with the degree of angiogenesis, but also with the levels of their counterparts in blood ^71^. Several other functions of PVMs have been described both in other healthy adult tissues (phagocytosis, antigen presentation, and immune regulation), and in the progression of pathological conditions (angiogenesis, metastasis, immunosuppression, recruitment of other leukocytes, and post-therapy relapse) ^55^. It is tempting to postulate that PVMs are critical not only for angiogenesis and hypervascularity in HCC through the secretion of VEGFA but are also responsible for the ‘leaky’ vessels seen in HCC that compromise immune control mechanisms. There is evidence from pre-clinical models to support both these possibilities. TIE2-expressing monocytes were found to be critical for tumour angiogenesis ^72^, supporting the fact monocyte-derived cells within tumours are key for angiogenesis. In addition, in a murine model of breast cancer, macrophage-specific depletion of VEGFA reduced both vascular permeability and circulating tumour cells whilst restoring vascular junctions ^53^.

Our TMA selection criteria of highly abundant TILs on H&E suggest we have selected areas that share features of the *Inflamed Class* of HCC ^70,73^. This class represents approximately 35 % of HCC tumours and is characterised by a microenvironment with increased interferon signalling and higher immune infiltration (increased TILs, macrophages, tertiary lymphoid structures, and elevated expression of checkpoint molecules) compared with non-inflamed tumours ^70^. The inflamed subclass exhibits a significantly higher fraction of CD8^+^ T cells and M1 macrophages when compared to the non-inflamed profiles. However, despite the presence of high TILs, these tumours persist. There could be many factors that contribute to this tumour persistence, including enrichment of clonally exhausted CD8^+^ T cells and regulatory T cells within HCC tissues ^68^. Our finding of no difference in the T cell density between tumour and non-tumour tissues is of interest. We also found that CD4^+^ T cells were more abundant than CD8^+^ T cells within the HCC TME, consistent with previous studies ^63^. Furthermore, CD4^+^ T cells expressing CD69 were the most common subset of CD4^+^ T cells across all regions in our study. Importantly, we found close interactions between these T cells and PVMs in the perivascular space. This niche could facilitate regulatory interactions for effector and resident T cells. We also identified multiple myeloid cells interacting with T cells within this niche, although the functional role of these myeloid cells remains unknown. Further studies are required to identify how and why T cells are potentially impaired within such tumour regions.

The findings from our study have several clinical implications. The identification of increased TIL density within a perivascular-immune neighbourhood (consisting of tumour endothelial cells and PVMs) warrants discussion of current therapeutic approaches that target immune-vascular cross-talk and the role of vascular normalisation in HCC ^74^. The tumour vasculature in HCC is structurally and functionally abnormal, and the tumour vessels are excessively leaky ^75^. Immune-vascular cross-talk underpins the rationale for using bevacizumab, an anti-VEGFA monoclonal antibody that normalises tumour vasculature, in conjunction with immune checkpoint inhibitors in advanced unresectable HCC ^5^. Advanced colorectal cancer patients receiving combined FOLFOX and bevacizumab have decreased VEGFB-mediated angiogenesis, including reduced myeloid cell-endothelial cell communication via VEGF as identified by scRNA-seq ^76^. In pre-clinical HCC models, anti-angiogenic therapy in conjunction with anti-PD-1 therapy further promotes vascular normalisation mediated by CD4^+^ T cells ^16^. Recently, biomarkers for response to systemic therapies in advanced HCC have been developed. Haber *et al.* found that tumours expressing high amounts of an 11-gene signature (IFNAP) captured a unique immune microenvironment characterised by a high gene expression of activated CD4^+^ memory T cells, M1 macrophages, and plasma cells. This novel signature predicted response and survival in patients treated in the first-line setting with anti-PD1 monotherapy ^77^. However, this study did not incorporate spatial analyses to determine immune cell interactions within the immune microenvironment captured by the IFNAP gene signature.

There are notable limitations to our study. Single-cell sequencing of TILs in HCC has identified eleven unique subsets: five CD8^+^ and six CD4^+^ T cell clusters including naïve, effector, and exhausted T cells, and mucosal-associated invariant T (MAIT) cells ^68^. We were unable to identify some of these subsets (undefined lymphocytes and undefined non-lymphocyte immune cells) due to constraints on the number of markers that can be included in a single IMC panel. Cell segmentation can become problematic when defining single cells sectioned from a three-dimensional organ. As such, cluster 44 was first identified as a single cell rather than two discrete closely interacting cells. This issue was resolved by examining representative images to confirm the presence of two discrete cells. Functionality of the PVM population was inferred by *in silico* analysis, rather than single cell analysis or cell isolation and culture experiments *ex vivo*. Finally, our sample size was small. Correlations with prognostic and clinicopathological data were performed but no associations were found, perhaps related to the limited sample size.

In summary, our study has identified a novel myeloid population in HCC characteristic of PVMs, and identified several immune cell and vascular neighbourhood interactions occurring more commonly within HCC tumours. Using IMC, we were able to comprehensively immunophenotype and spatially map the immune landscape of human HCC tumour and adjacent non-tumour tissue. Further studies are required to evaluate the function of these immune cells, their effect on immune-mediated tumour control and therapeutic responses, and their role in hepatocarcinogenesis and HCC progression.

## Acknowledgements

Assistance with the study: We gratefully acknowledge the subsidised access to Sydney Cytometry and thank the support staff in this core facility for their assistance. We thank the Australian Centre for Microscopy & Microanalysis at the University of Sydney for their assistance with this work.

Presentations: Preliminary data were presented at the 2023 EASL Liver Cancer Summit (Estoril, Portugal); 2023 CYTO (Montreal, Canada); and GESA AGW (Brisbane, Australia).

## List of abbreviations

FFPE: Formalin-fixed paraffin embedded
H&E: Haematoxylin and eosin
HCC: Hepatocellular carcinoma
IHC: Immunohistochemistry
IMC: Imaging mass cytometry
LSEC: Liver sinusoidal endothelial cell
MAIT: cells mucosal-associated invariant T cells
mIHC: Multiplex immunohistochemistry
MRS: myeloid response score
MVI: Microvascular invasion
PCA: Principal component analysis
PERMANOVA: Permutational multivariate analysis of variance
PVM: perivascular macrophage
ROI: Region of interest
RT: Room temperature
scRNA-seq: Single-cell RNA-sequencing
sPLS-DA: Sparse partial least squares-discriminant analysis
TAM: tumour-associated macrophage
TGF-β: transforming growth factor-beta
TIL: Tumour-infiltrating lymphocyte
TMA: Tissue microarray
TME: Tumour immune microenvironment
TNM: Tumour-node-metastasis
TSA: Tyramide signalling amplification
VEGFA: Vascular endothelial growth factor A

## Data availability

Data are available on Zenodo (DOI 10.5281/zenodo.10547481).

## Supplementary figure legends

**Supplementary Figure 1.**
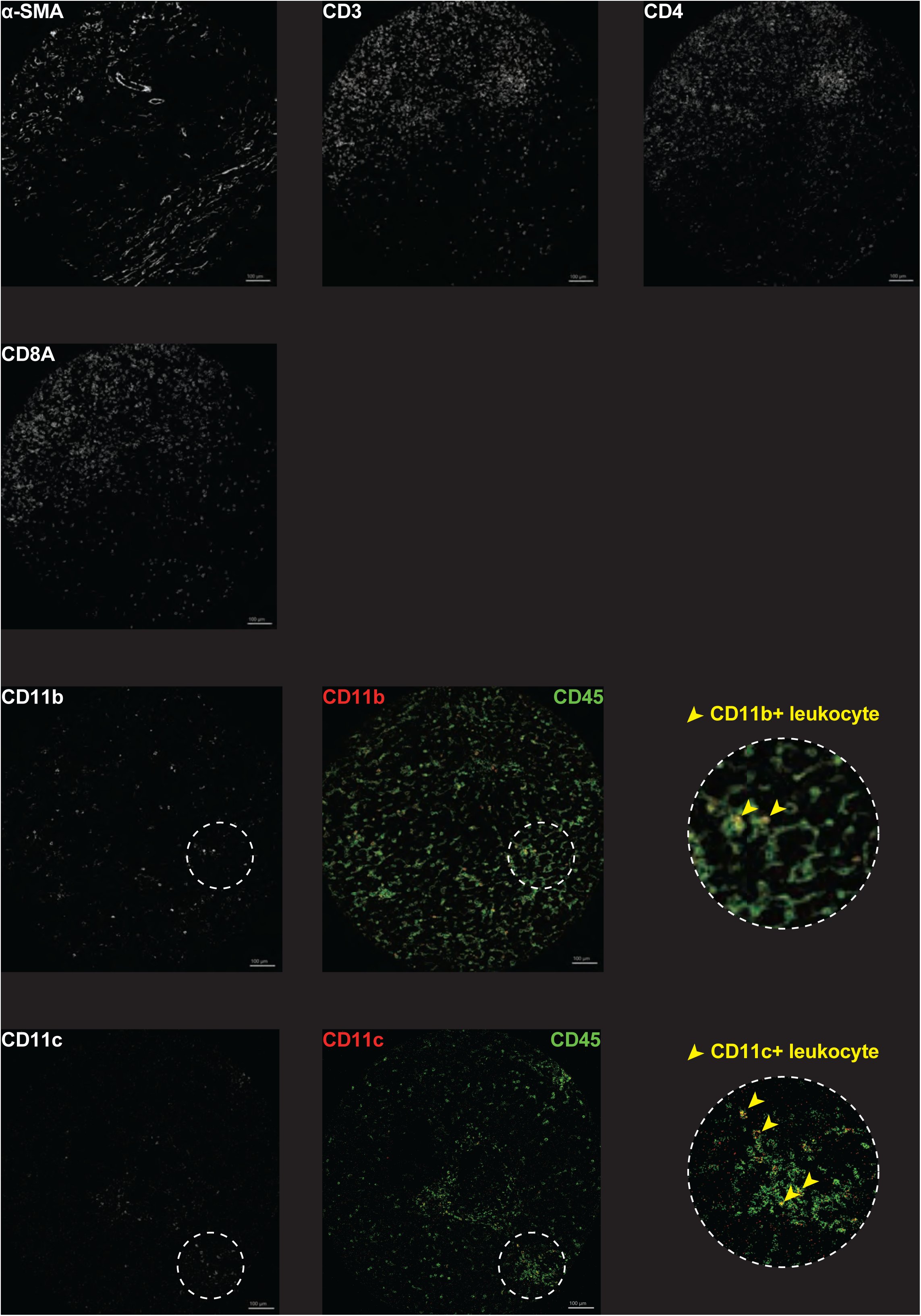
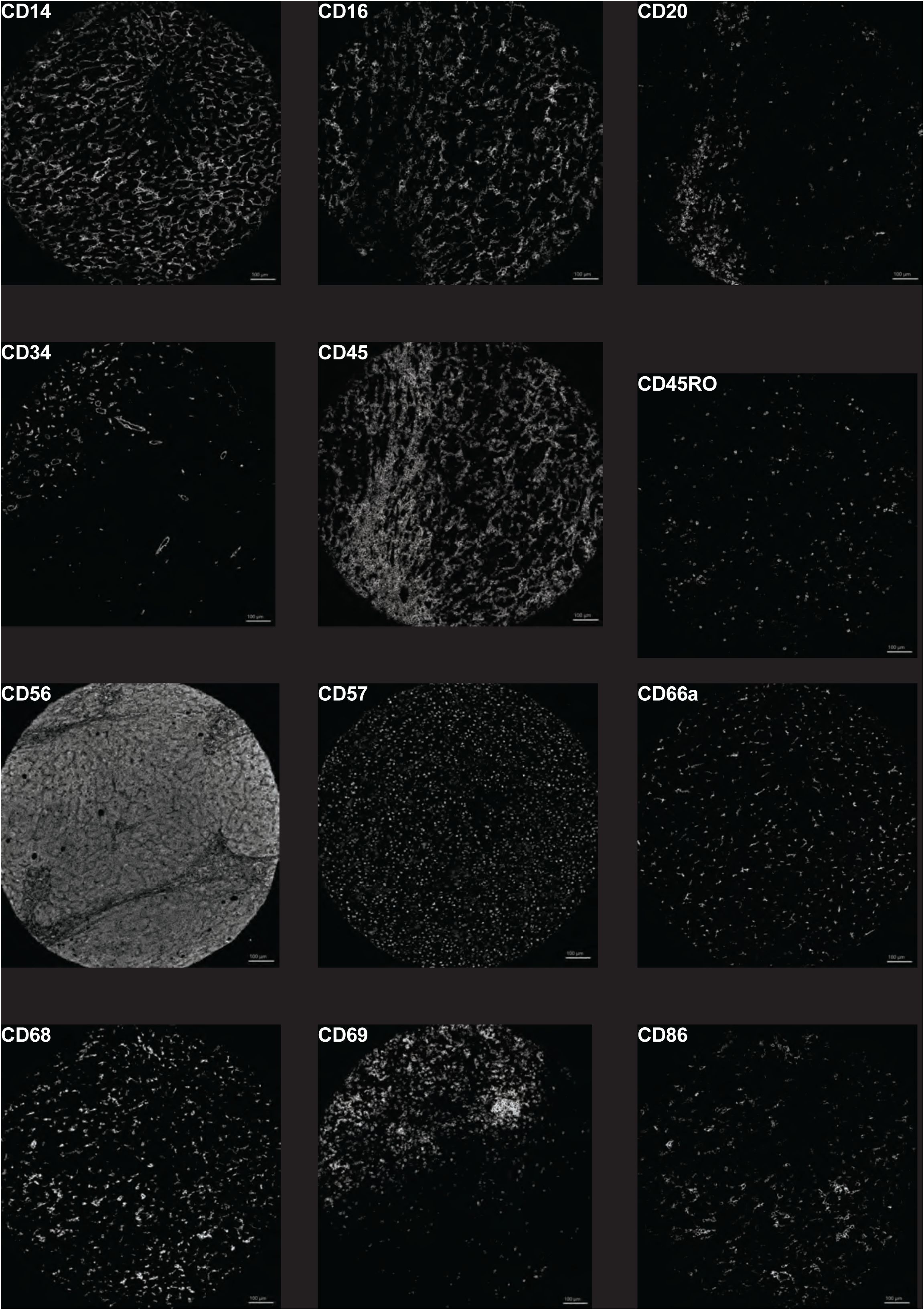
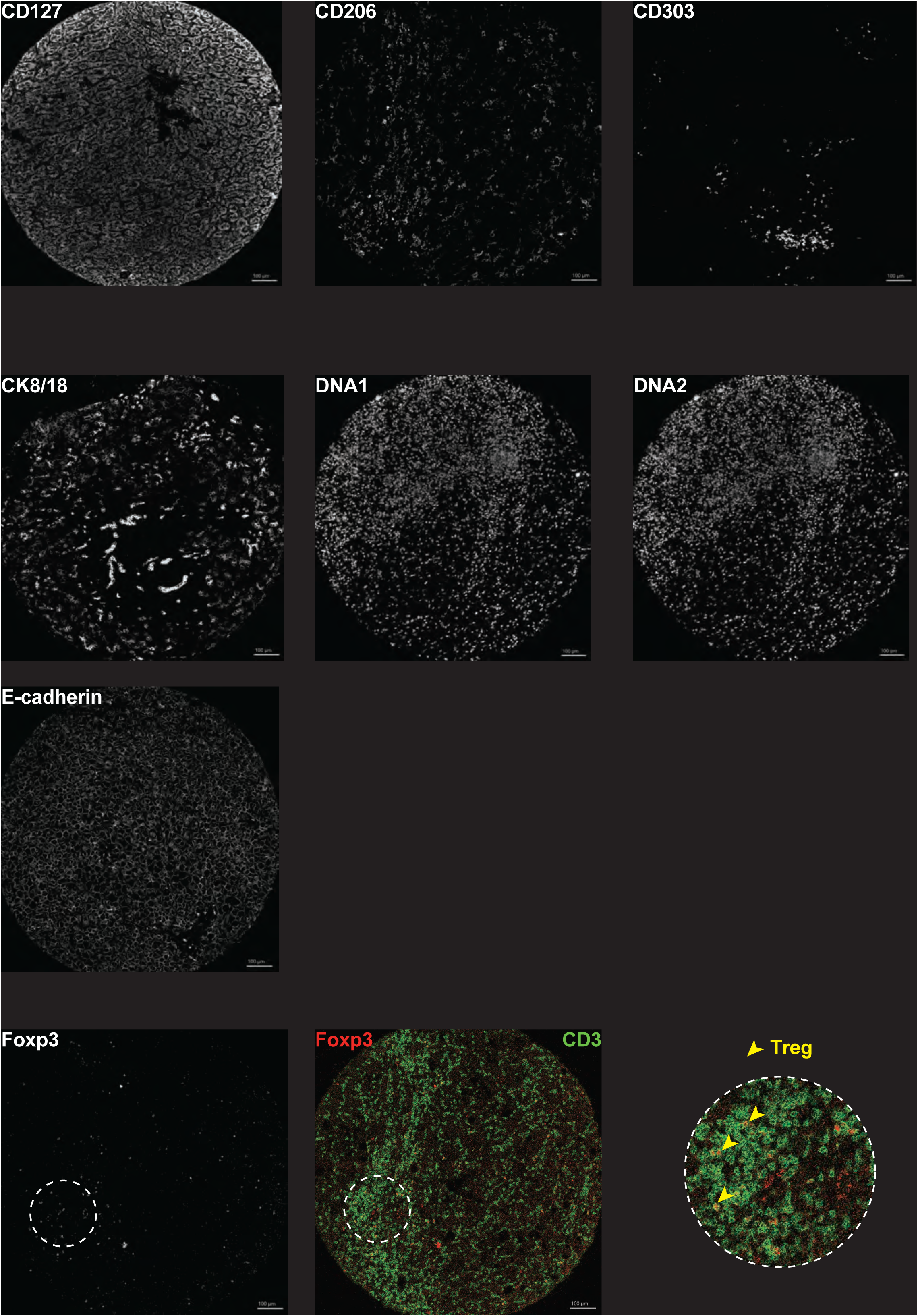
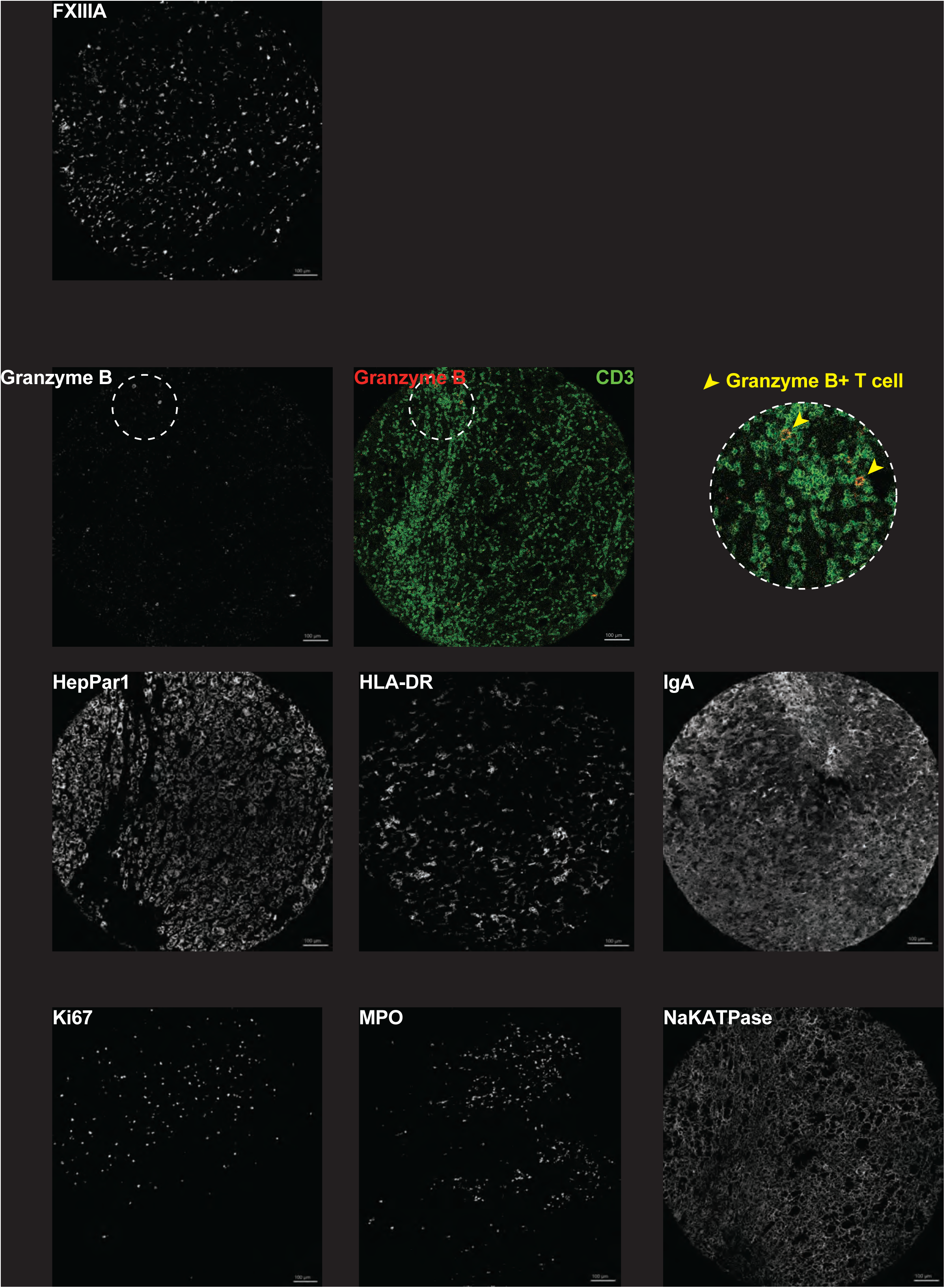
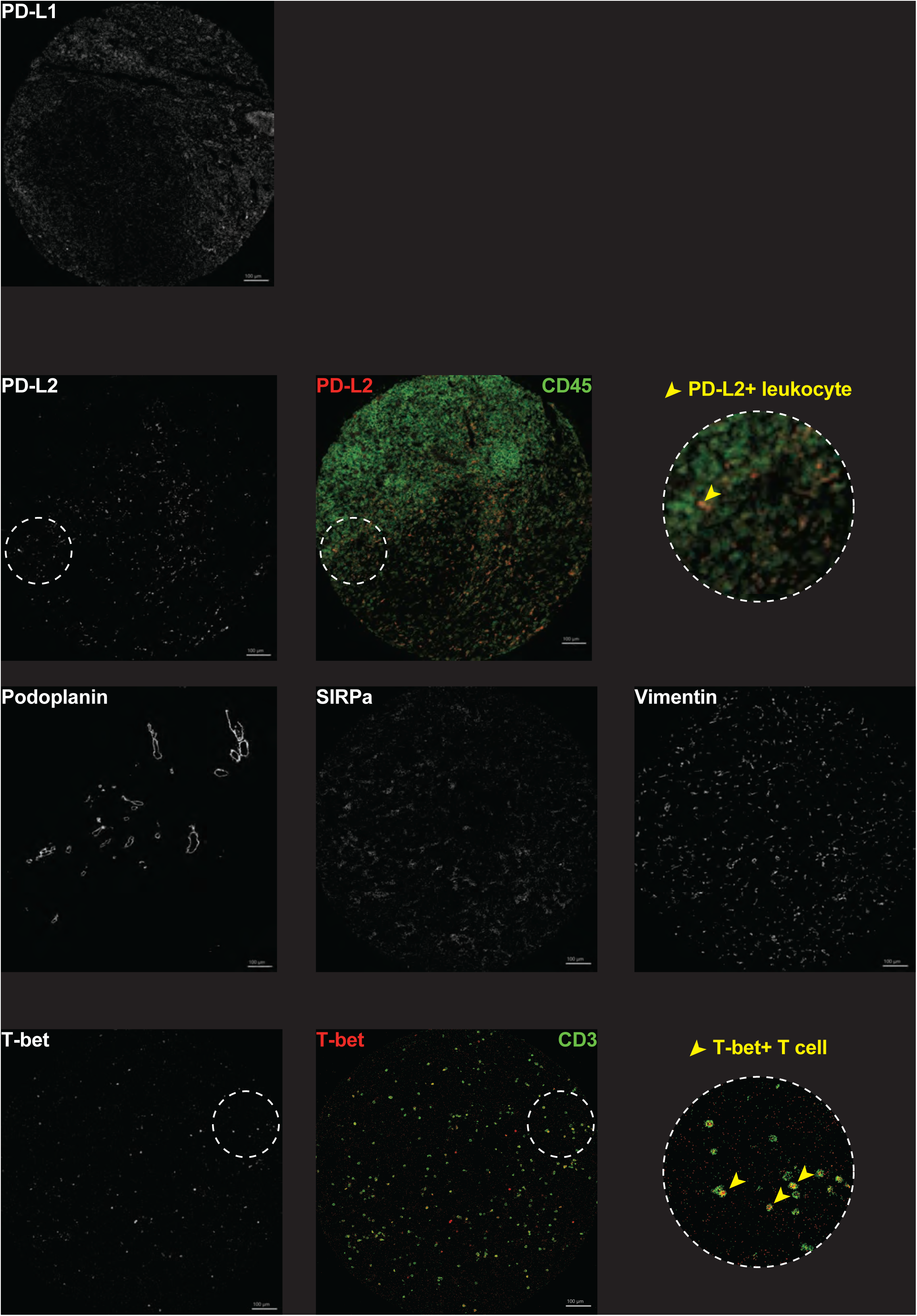
Single marker IMC staining. Representative images for individual IMC antibodies.

**Supplementary Figure 2.**
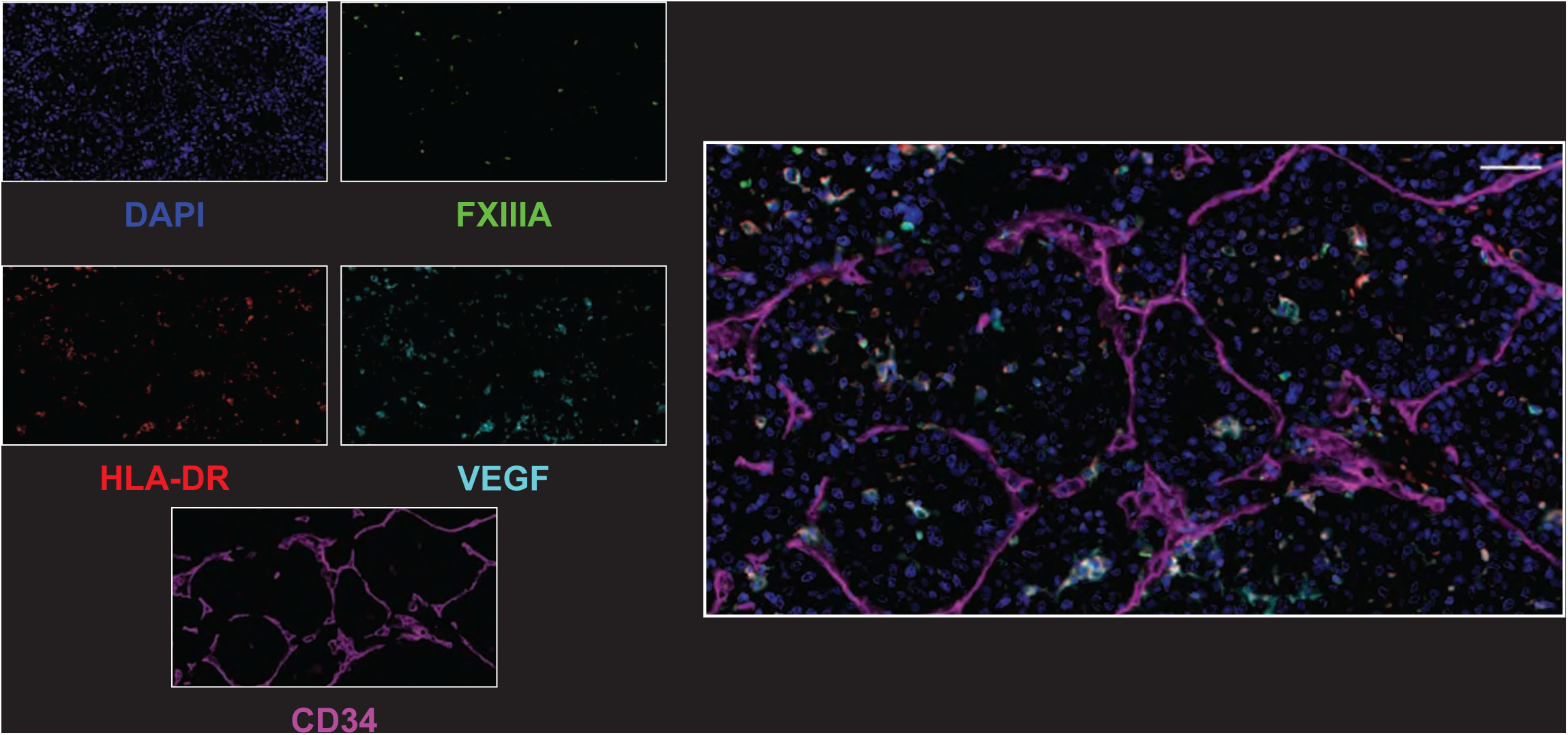
Single marker OPAL staining. Representative images for individual OPAL antibodies.

**Supplementary Figure 3.**
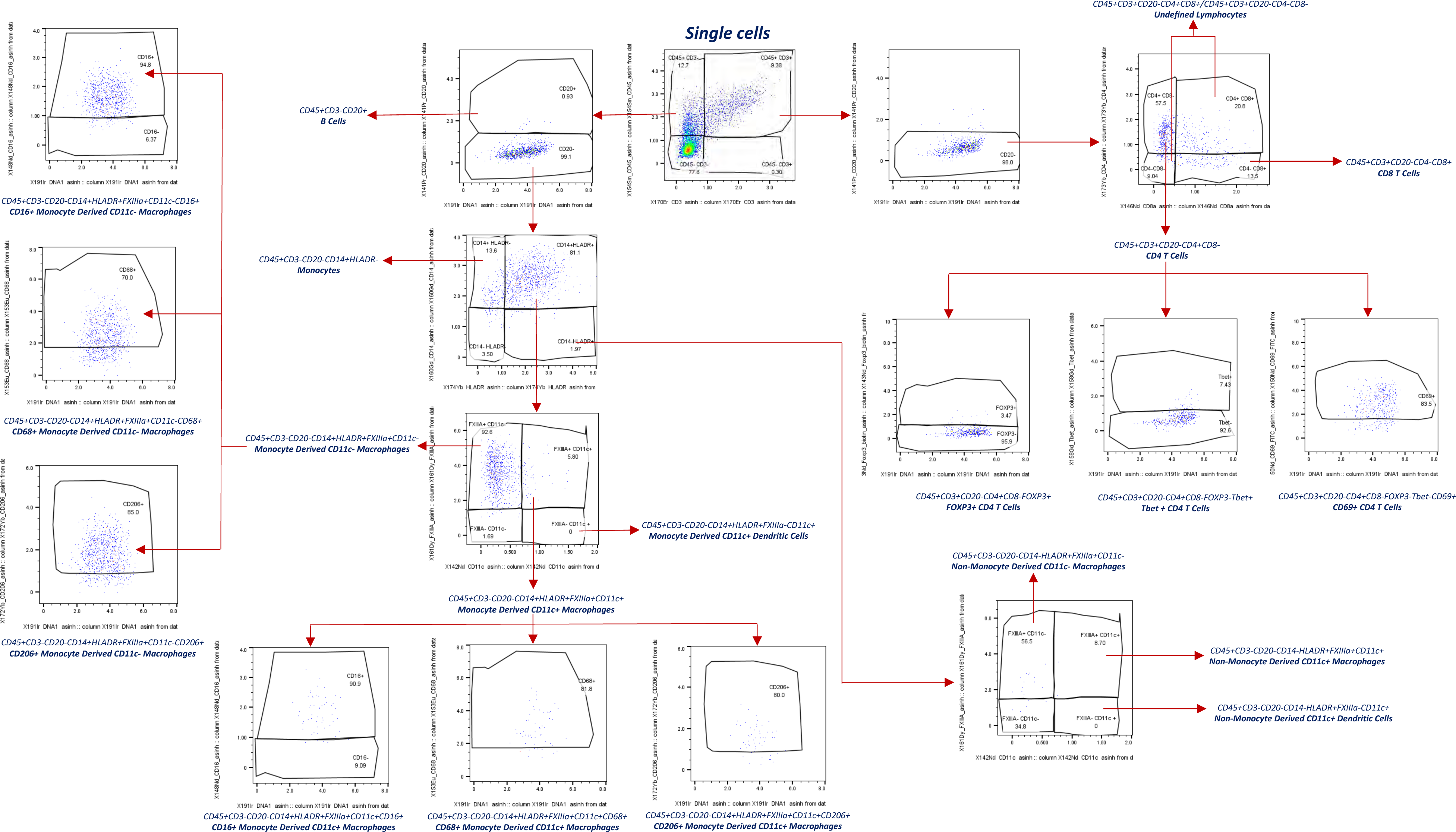
Manual gating strategy of immune cell subsets. Representative gating strategy identifying immune cell subsets.

**Supplementary Figure 4.**
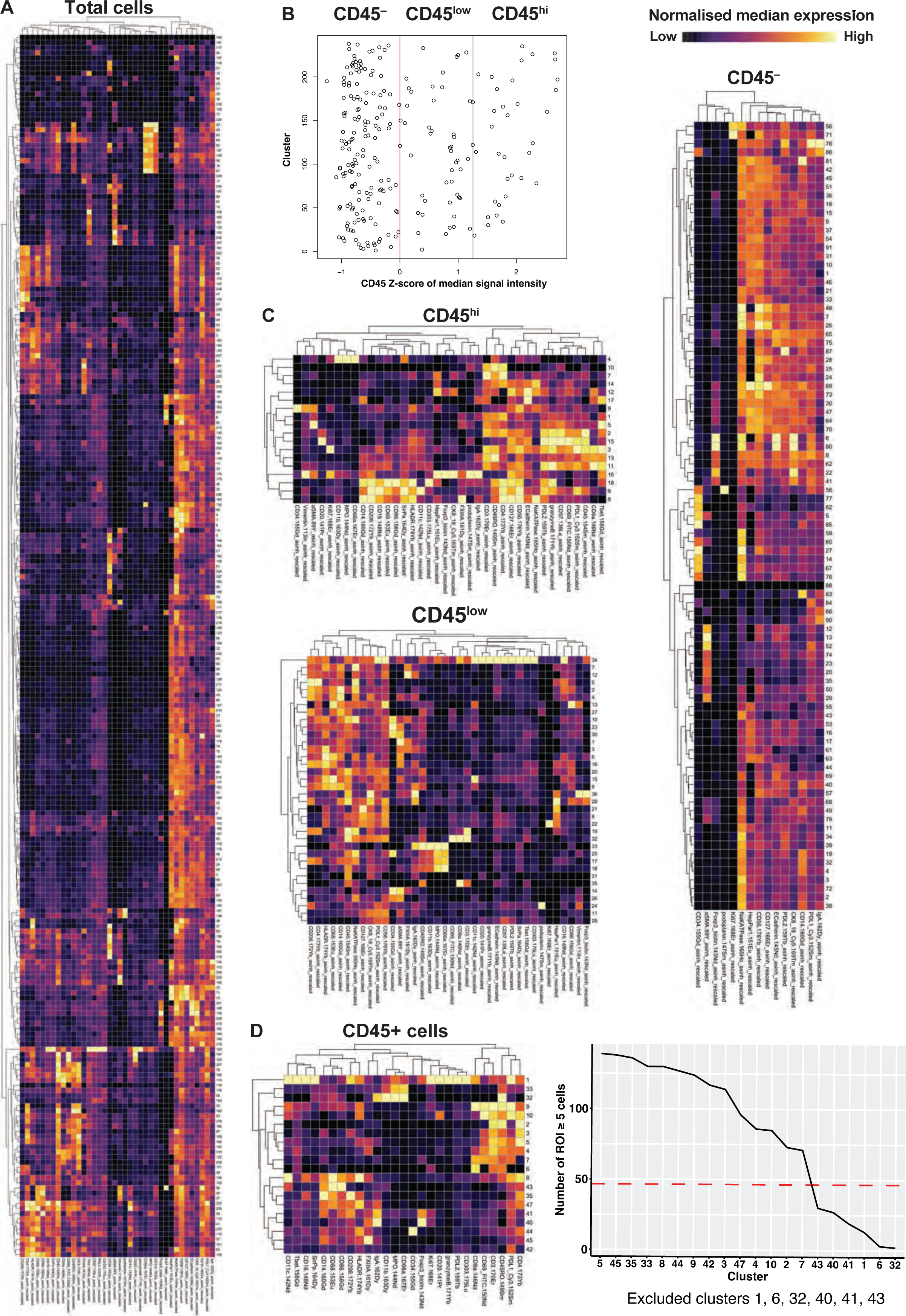
Clustering of single cells. A) Total cells were clustered using X-shift. B) The median signal intensity of CD45 was calculated for each cluster before running z-score normalisation. Red vertical line is at zero, whereby a negative cluster indicates no CD45 expression, whilst a positive value indicates CD45 expression. The blue line represents the absolute value of the smallest value, used to differentiate between CD45^low^ and CD45^hi^ populations. C) Cells then underwent X-shift clustering within each of the three groups (CD45^−^, CD45^low^, CD45^hi^) resulting in 91, 36, and 18 clusters respectively. Similar clusters were then combined using the function simprof as part of the ‘clustsig’ R package, generating 47 clusters (20 CD45^+^ clusters and 27 CD45^−^ clusters). D) The number of CD45^+^ clusters with ≥5 cells in each region were calculated. Clusters with <5 cells across less than a third of total regions were excluded, which removed six clusters. Heatmaps show the scaled median signal expression of each marker (columns) across each cluster (rows).

**Supplementary Figure 5.**
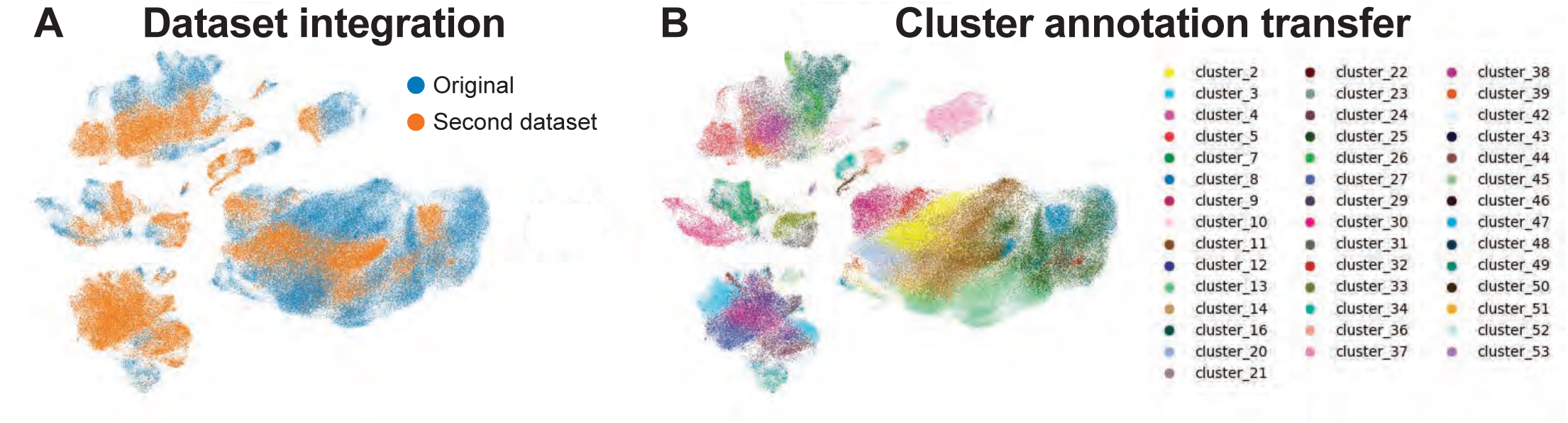
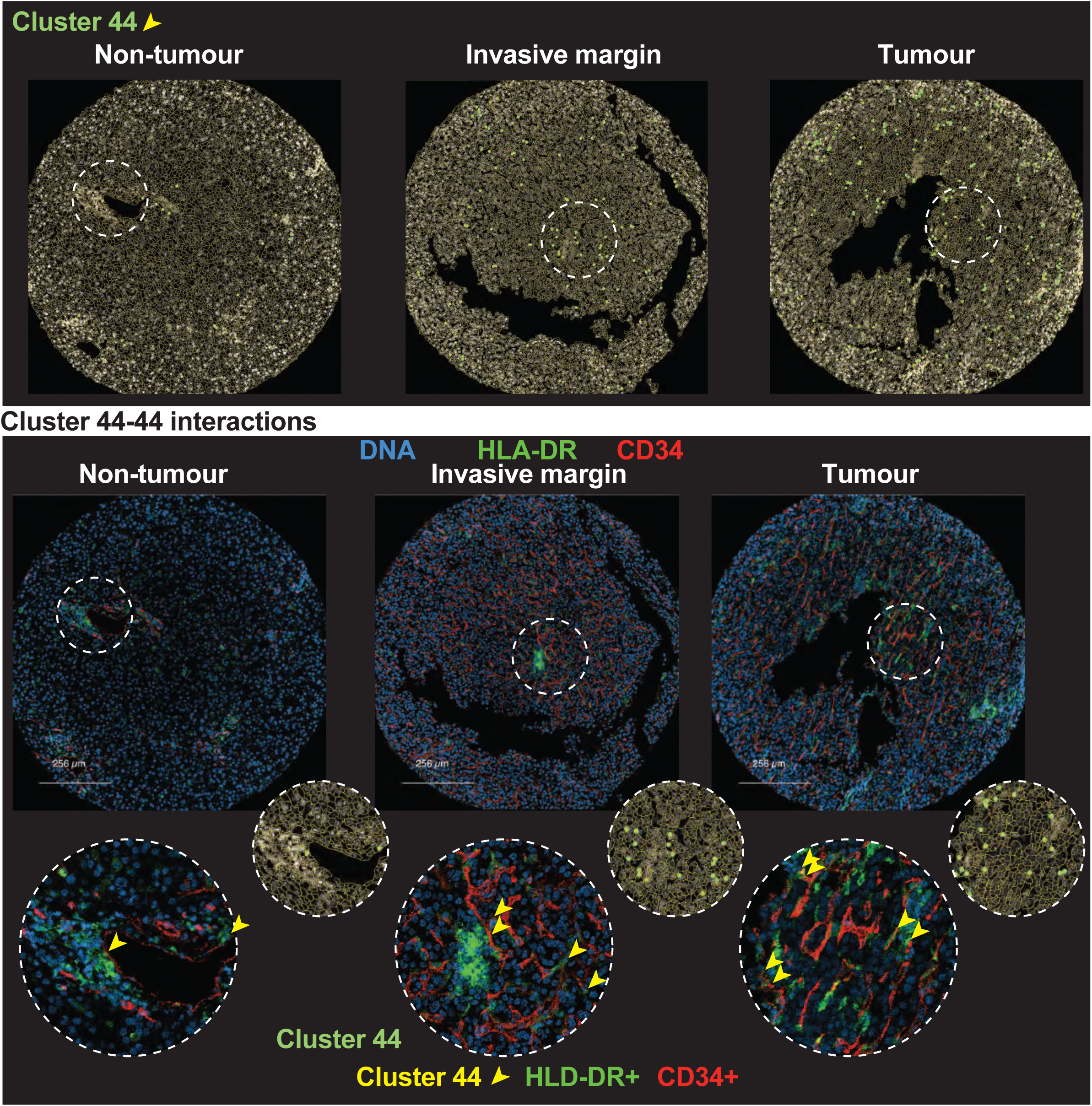
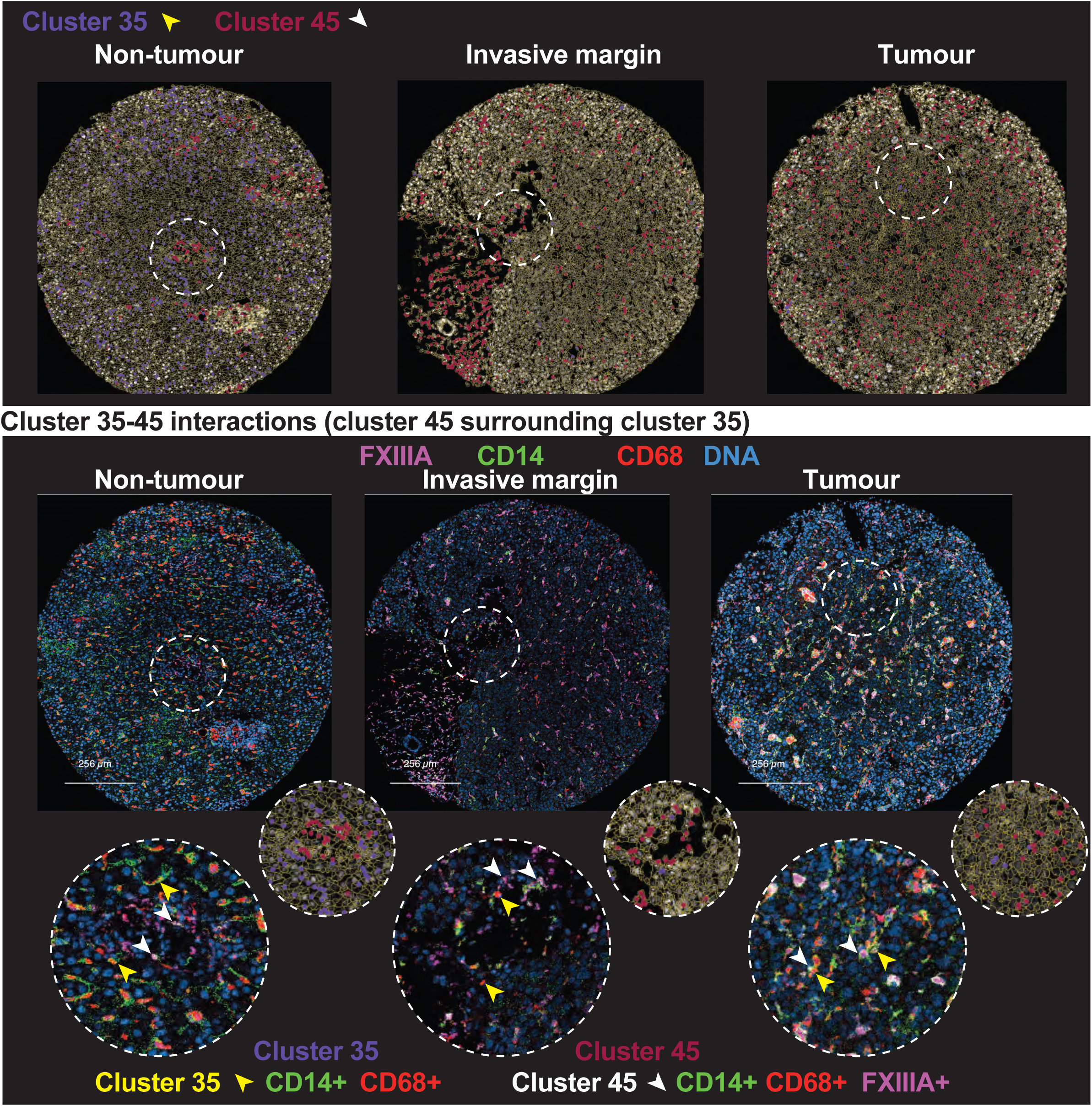
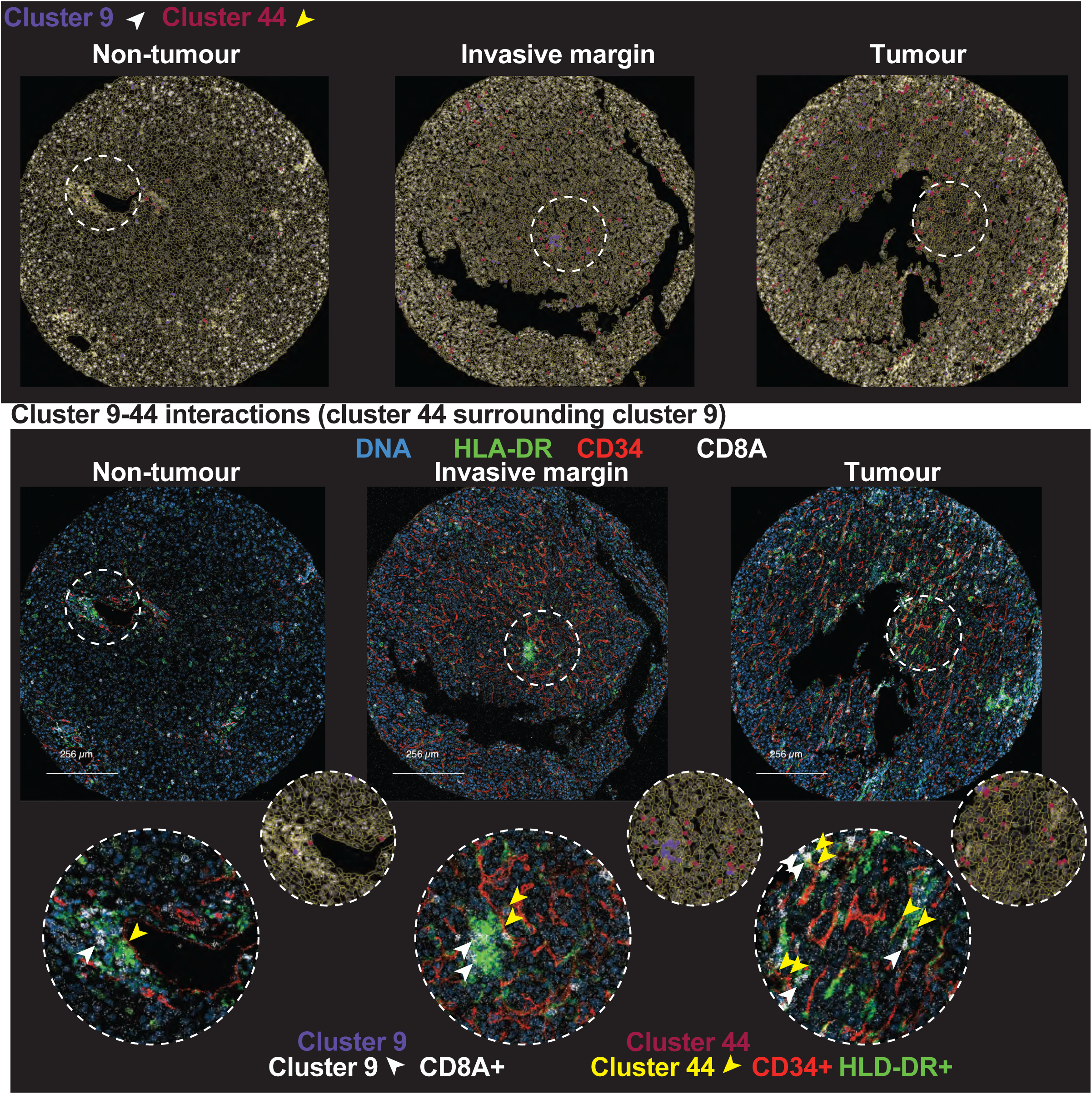
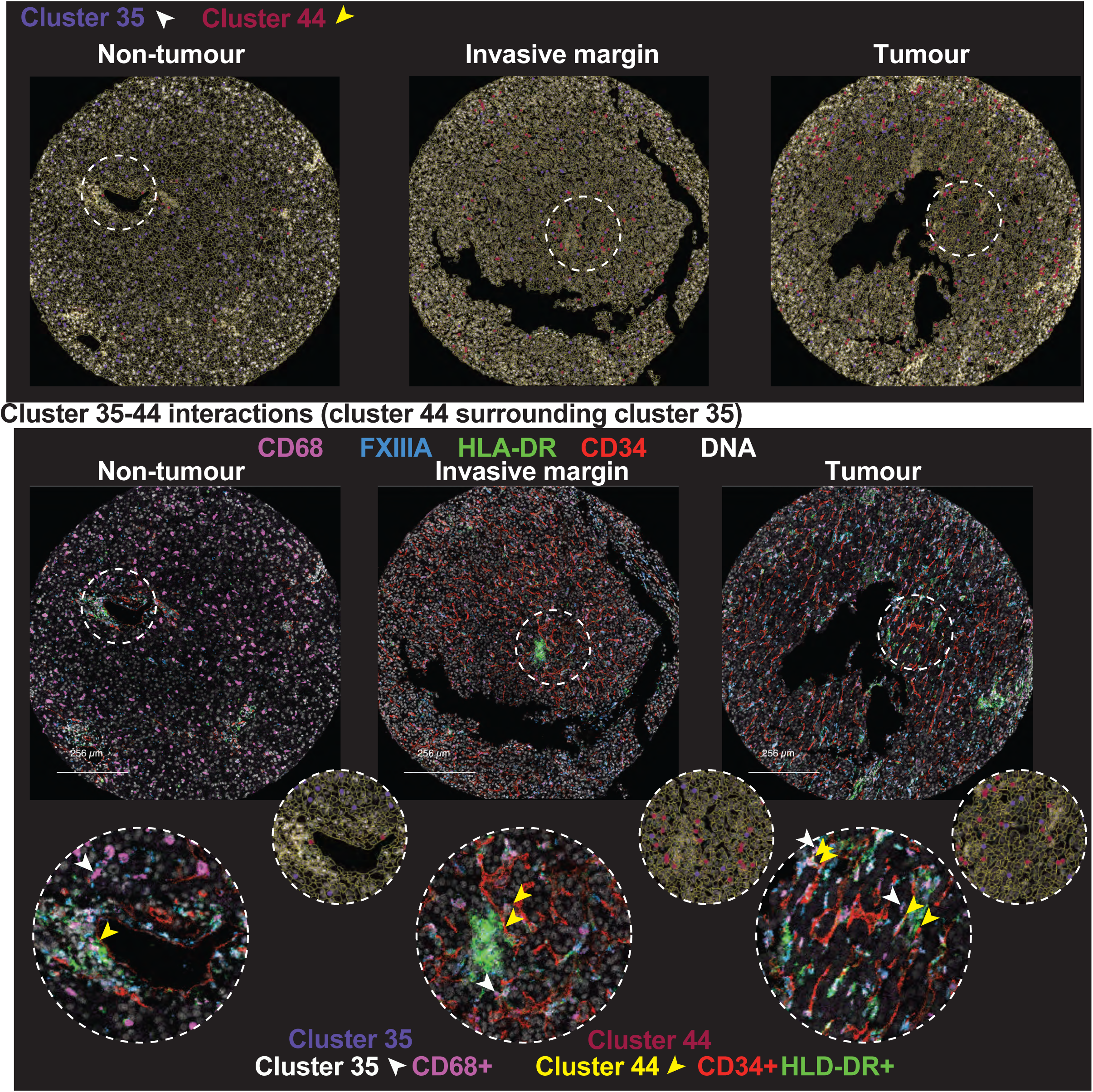
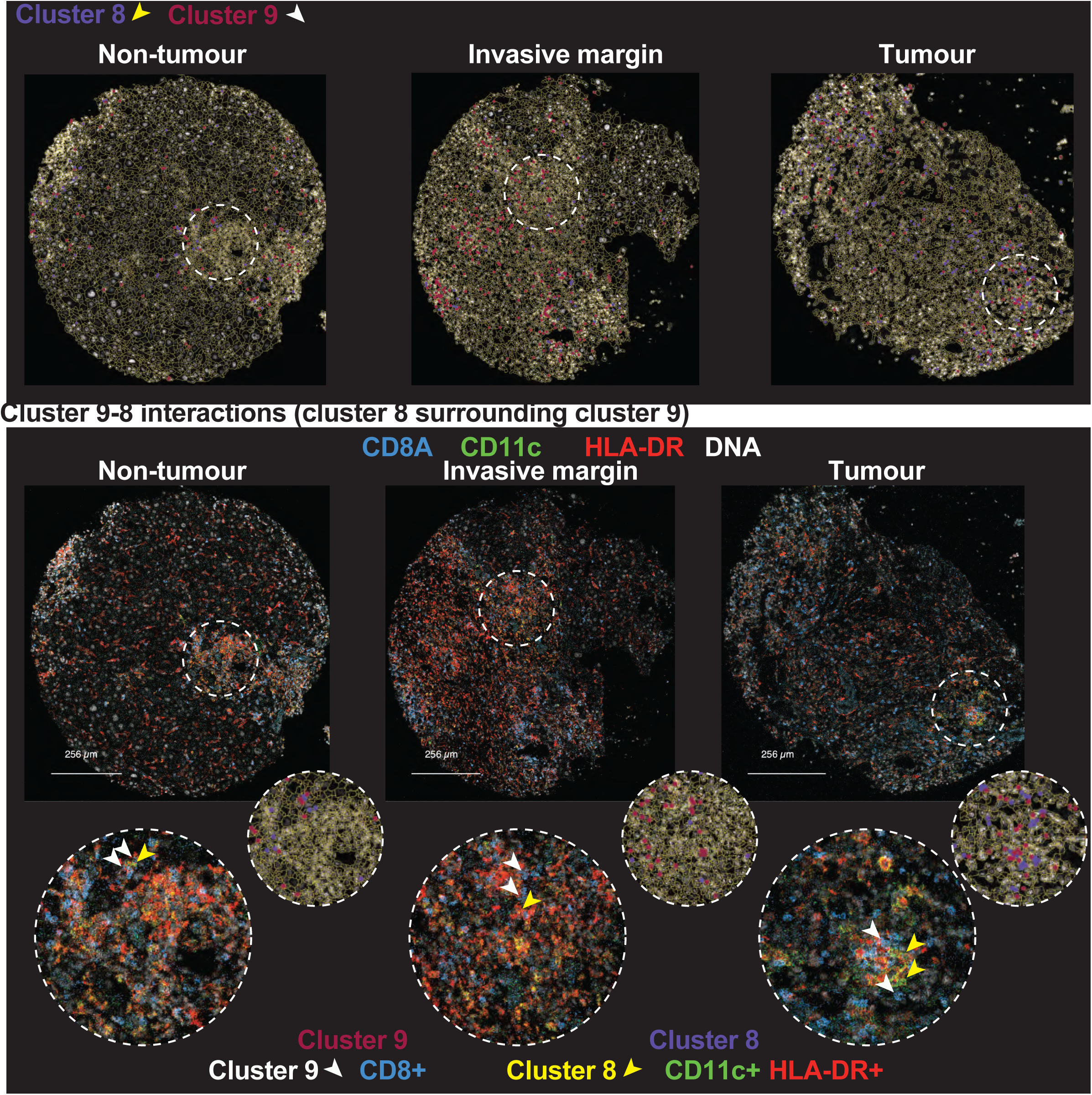
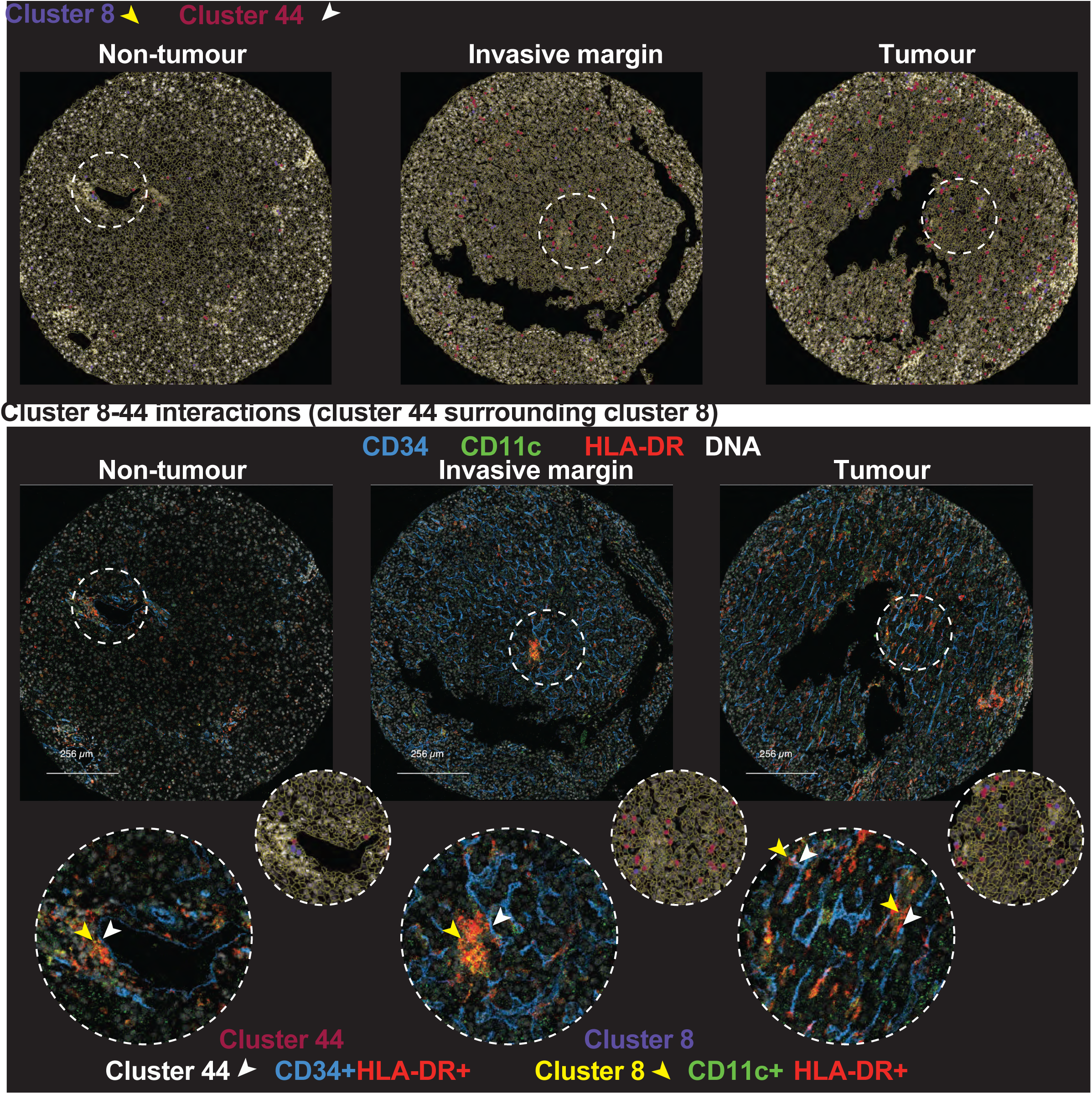
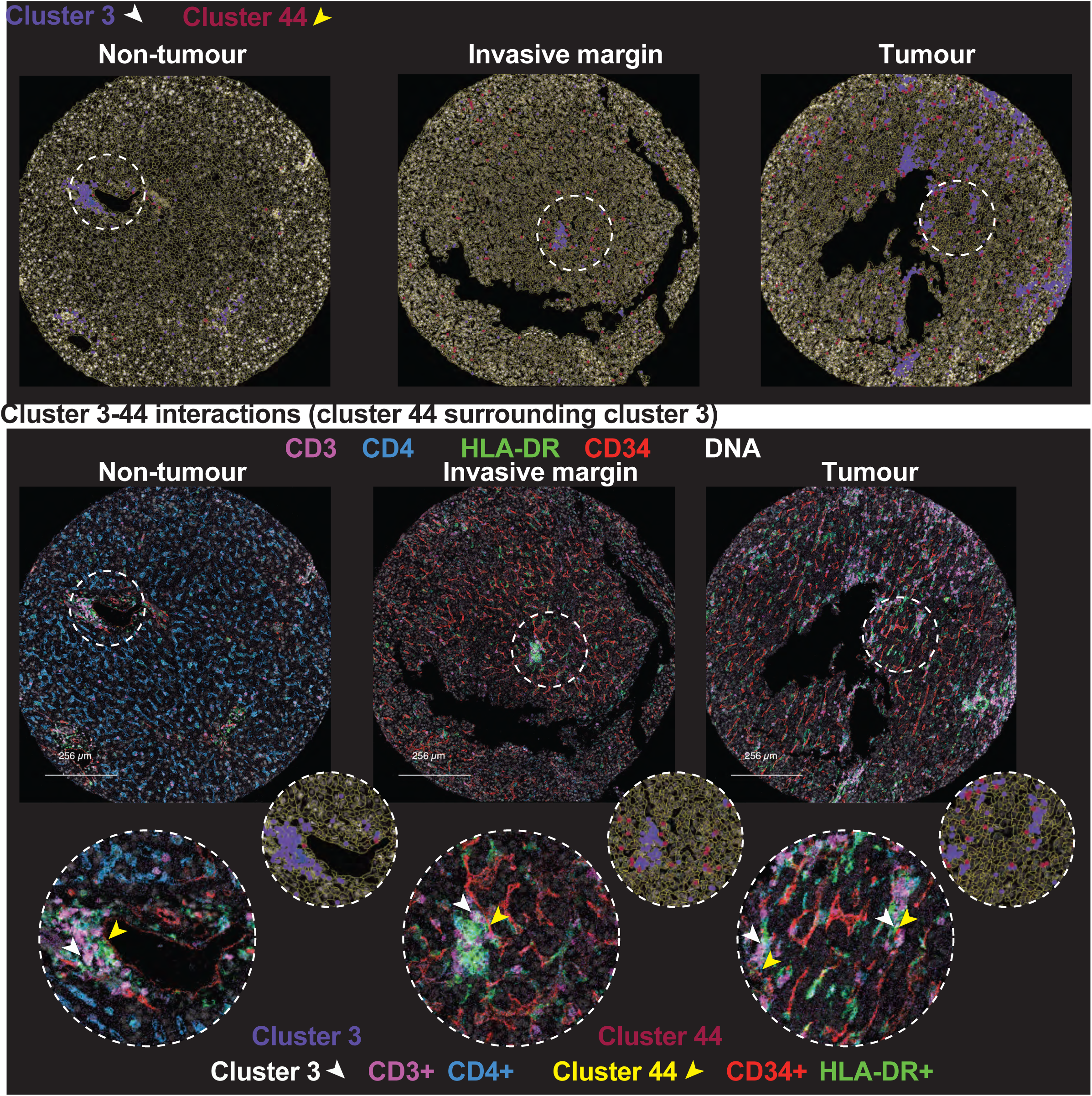
scANVI cluster annotation transfer. Cluster annotations were transferred from initial single-cell RNA-sequencing (scRNA-seq) dataset (GSE149614) to a second scRNA-seq dataset (https://data.mendeley.com/datasets/skrx2fz79n/1). A) UMAP visualising the integrated original reference dataset (orange) with the second dataset (blue). B) UMAP of predicted annotated clusters.

**Supplementary Figure 6.**
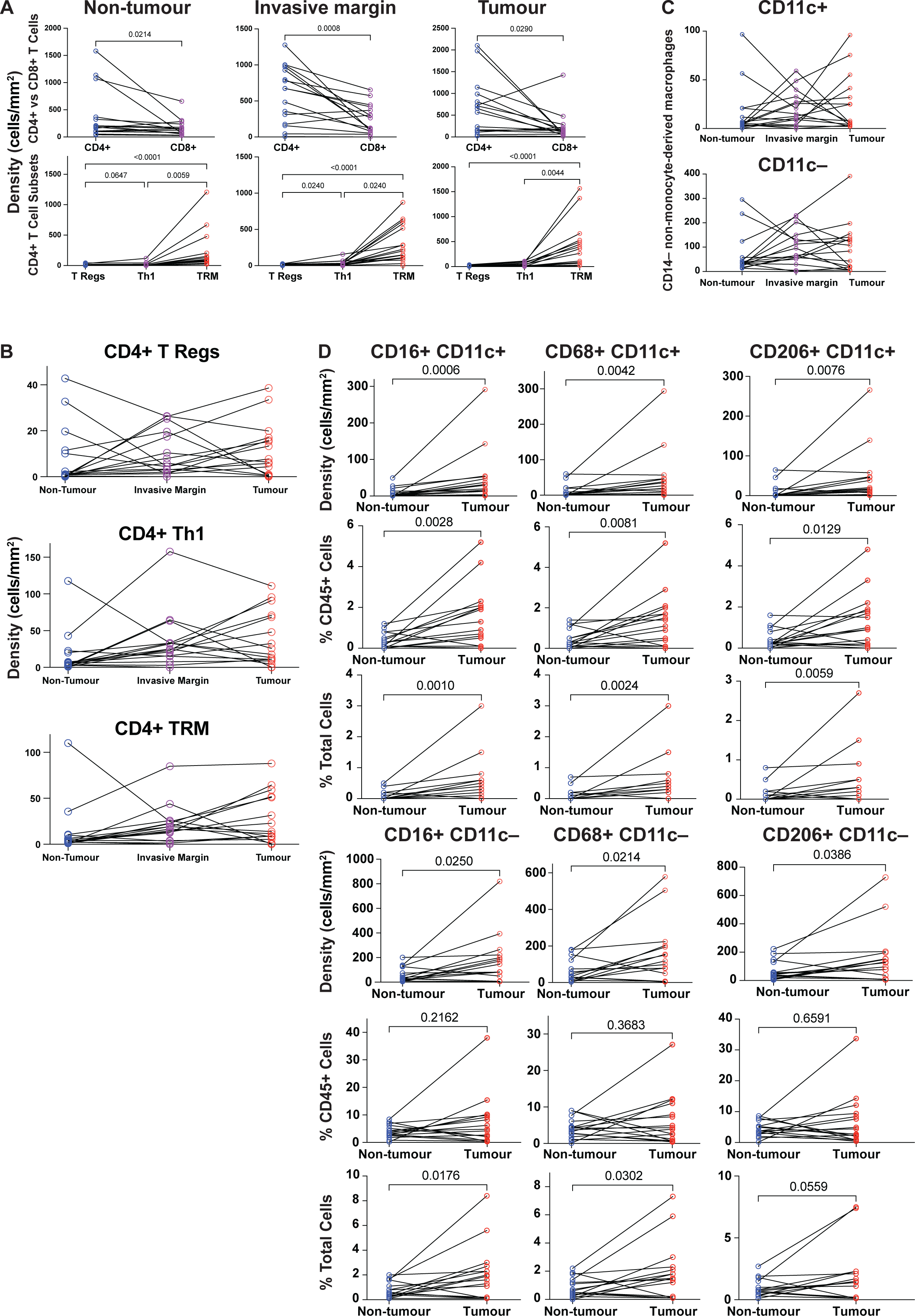
T cell and myeloid cell subset levels across regions. A) Densities of T cell subsets across regions. B) Densities of CD4^+^ T cell subsets. Friedman test with Dunn’s multiple comparison corrections was done. C) Densities of CD14^−^ non-monocyte-derived macrophages across regions. D) Density and proportions (% of CD45^+^ cells or total cells) across myeloid cell subsets. Wilcoxon test was used.

**Supplementary Figure 7.**
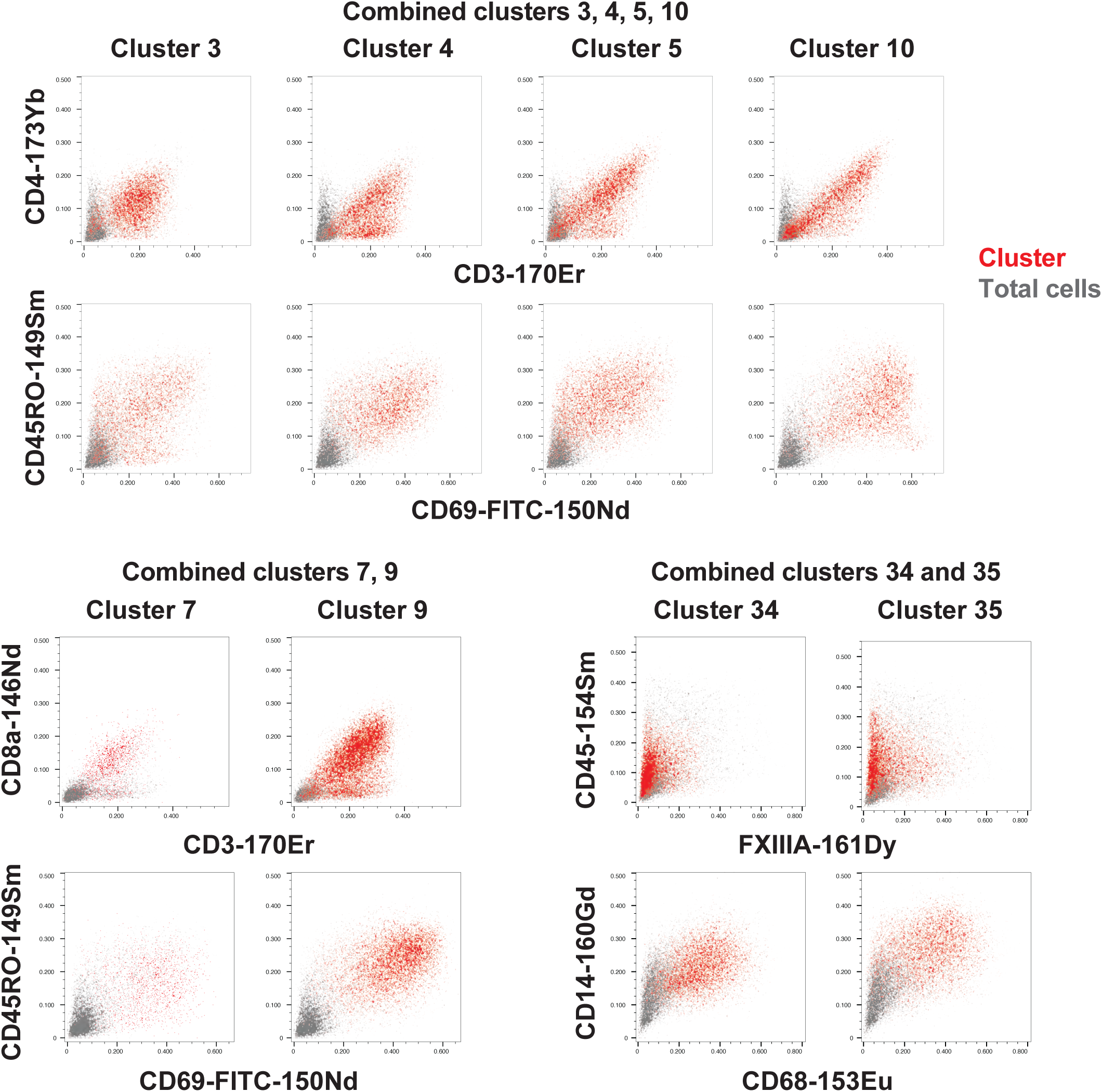
Phenotypically similar clusters were combined. CD45^+^ clusters that were phenotypically similar were combined. Representative images show each cluster (red) compared to total cells (grey).

**Supplementary Figure 8.** Representative images of parameters that contributed to differences between non-tumour and tumour regions. Representative images of cluster results on segmented data (green, dark blue, dark red dots). Images of imaging mass cytometry with markers and clusters as indicated.

**Supplementary Figure 9.**
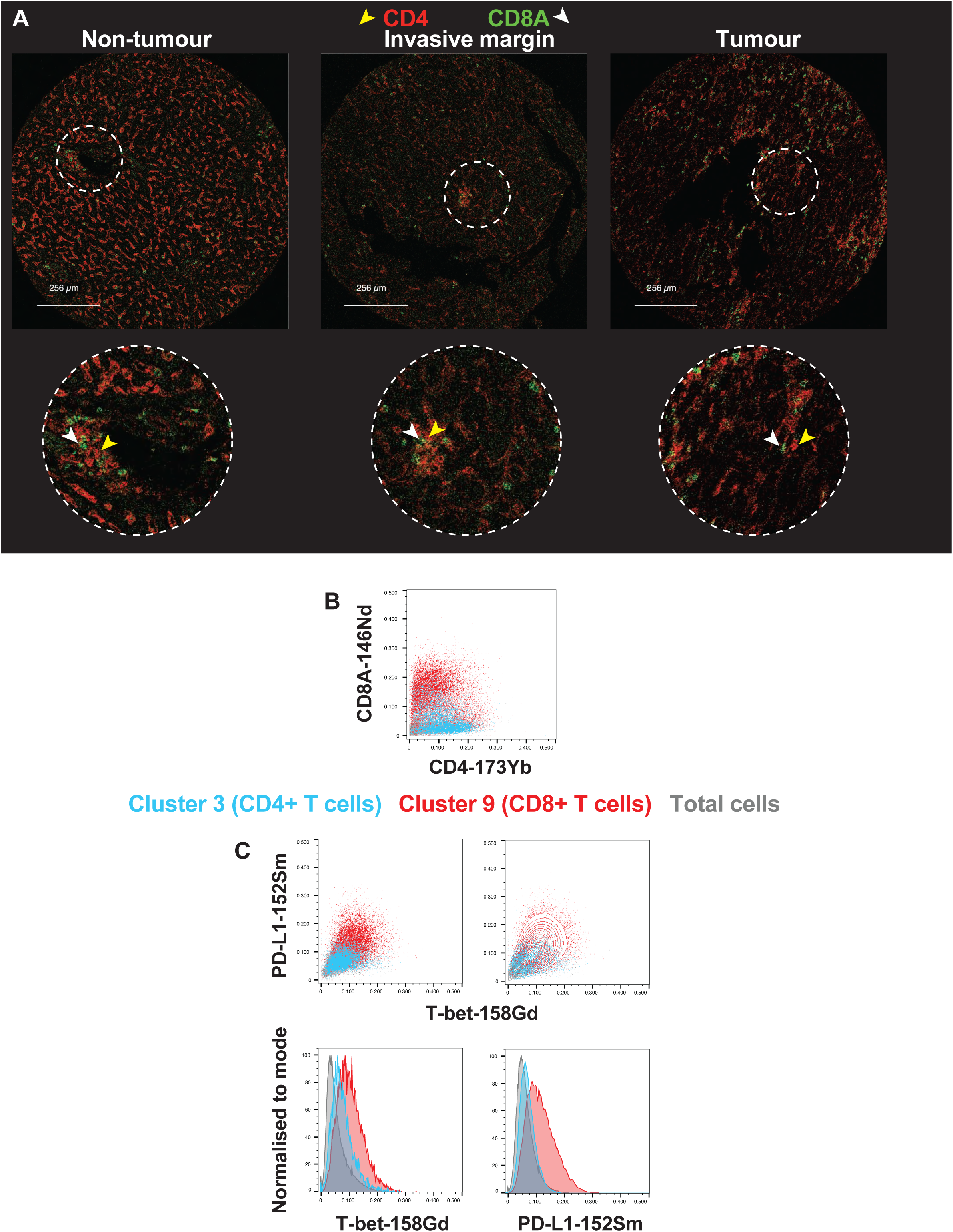
T cell marker expression. A) Representative image of CD4 (red) and CD8A (green) staining. B) Plot showing cluster 3 (blue, CD4^+^ T cells), cluster 9 (red, CD8^+^ T cells), and total cells (grey) against CD4 and CD8 staining. C) Plots showing cluster 3 (blue, CD4^+^ T cells), cluster 9 (red, CD8^+^ T cells), and total cells (grey) for PD-L1 and T-bet expression.

**Supplementary Figure 10.**
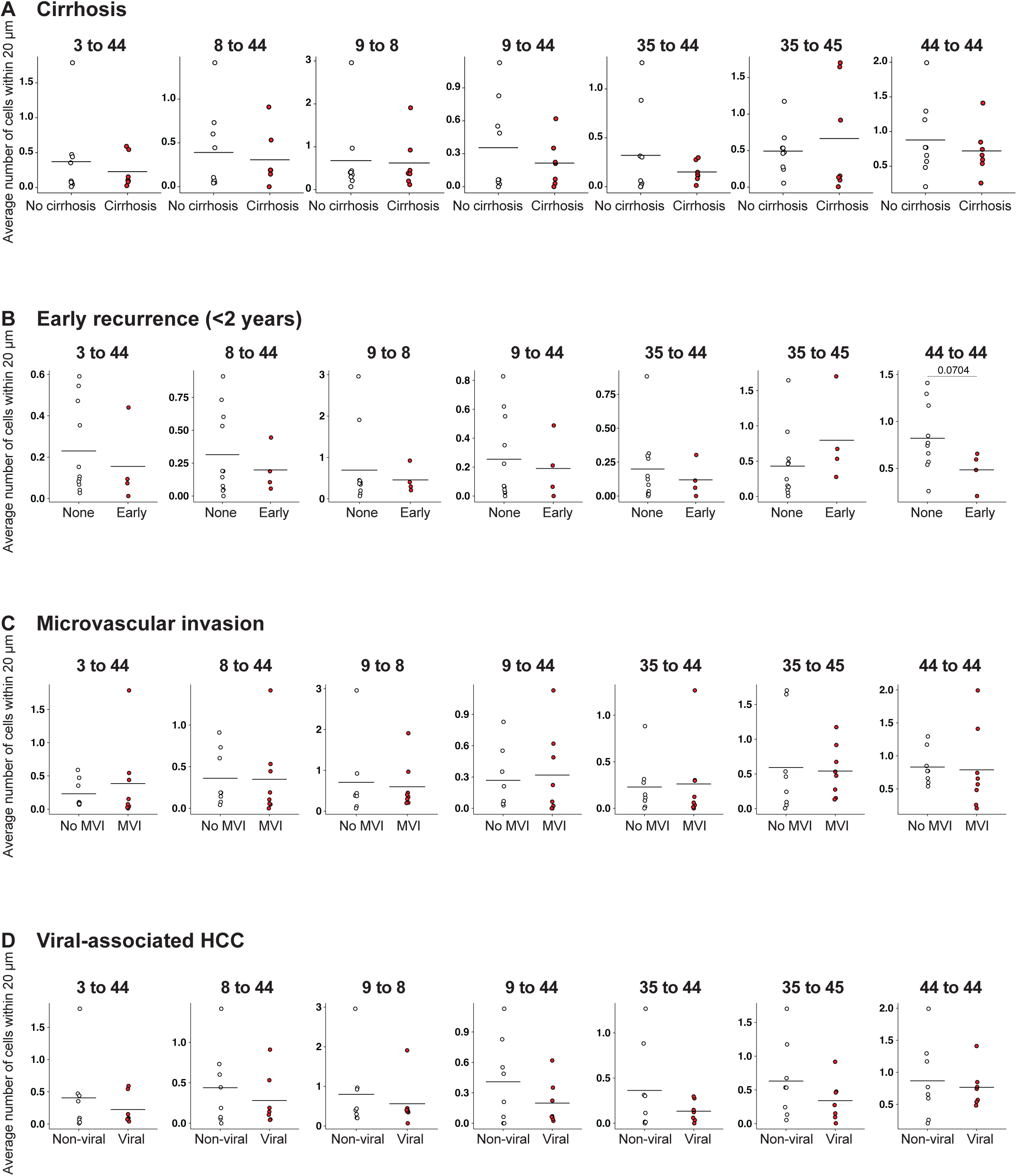
No association between clinical parameters and tumour neighbourhood interaction. Tumour neighbourhood interactions were correlated with A) cirrhosis, B) early recurrence (<2 years), C) microvascular invasion (MVI), and D) viral-associated HCC. Permutation student’s t-test.

**Supplemenary Table 1:** Patient cohort.

| ID | Experiment | Aetiology | Cirrhotic | BCLC | TNM | Largest tumour size (mm) | Tumour number | MVI | Differentiation | Previous Tx |
| --- | --- | --- | --- | --- | --- | --- | --- | --- | --- | --- |
| 1 | IMC (TMA1) | HBV | Y | A | pT2 | 17 | 2 | Y | Moderate | N |
| 2 | IMC (TMA1) | MASH | N | A | pT2 | 165 | 1 | Y | Moderate | N |
| 3 | IMC (TMA1) | HBV | Y | A | pT1 | 40 | 2 | N | Well | N |
| 4 | IMC (TMA1) | HCV, MASH | Y | A | pT1b | 32 | 1 | N | Poor | N |
| 5 | IMC (TMA1) | ALD | Y | A | T1bN0MX | 35 | 1 | N | Moderate | N |
| 6 | IMC (TMA1) | MASH | N | A | pT1b | 37 | 1 | N | Moderate | N |
| 7 | IMC (TMA1) | MASH | N | A | pT3 | 270 | 1 | Y | Moderate | N |
| 8 | IMC (TMA1) | HBV | N | A | pT2 | 117 | 1 | Y | Well to moderate | N |
| 9 | IMC (TMA2) | HCV | Y | A | pT2 | 30 | 1 | Y | Moderate | N |
| 10 | IMC (TMA2) | HCV | N | A | pT2 Nx | 170 | 1 | Y | Moderate | N |
| 11 | IMC (TMA2) | ALD | Y | A | pT2 | 25 | 1 | Y | Well to moderate | N |
| 12 | IMC (TMA2) | Unknown | N | B | T3 N0 M0 | 110 | >3 | Y | Moderate | N |
| 13 | IMC (TMA2) | HCV | Y | A | pT2 | 21 | 1 | N | Well to moderate | N |
| 14 | IMC (TMA2) | HBV | N | A | pT1a | 16 | 1 | N | Moderate | N |
| 15 | IMC (TMA2) | MASH, ALD | N | A | pT1a | 9 | 1 | N | Well | N |
| 16 | IMC (TMA2) | Unknown | N | A | pT1b N0 | 140 | 1 | N | Poor | N |
| 17 | OPAL | HBV | N | A | pT1b N0M0 | 150 | 1 | Y | Moderate | N |
| 18 | OPAL | Unknown | Y | A | pT1b N0M0 | 120 | 1 | N | Well | TACE |
| 19 | OPAL | ALD | N | A | pT2 N0M0 | 130 | 1 | Y | Moderate | N |
| 20 | OPAL | HCV, ALD | Y | B | pT2 N0M0 | 45 + 18 | 2 | N | Moderate | N |
| 21 | OPAL | Unknown | N | A | pT2 N0M0 | 160 | 1 | Y | Poor | Atezolizumab |
| 22 | OPAL | HCV | N | A | pT4 N1 M0 | 45 | 1 | Y | Poor | N |
| 23 | OPAL | Unknown | N | A | pT2 N0M0 | 180 | 1 | Y | Moderate | N |
| 24 | OPAL | HBV | N | A | pT1b N0M0 | 71 | 1 | N | Moderate | N |

**Supplementary Table 2:**
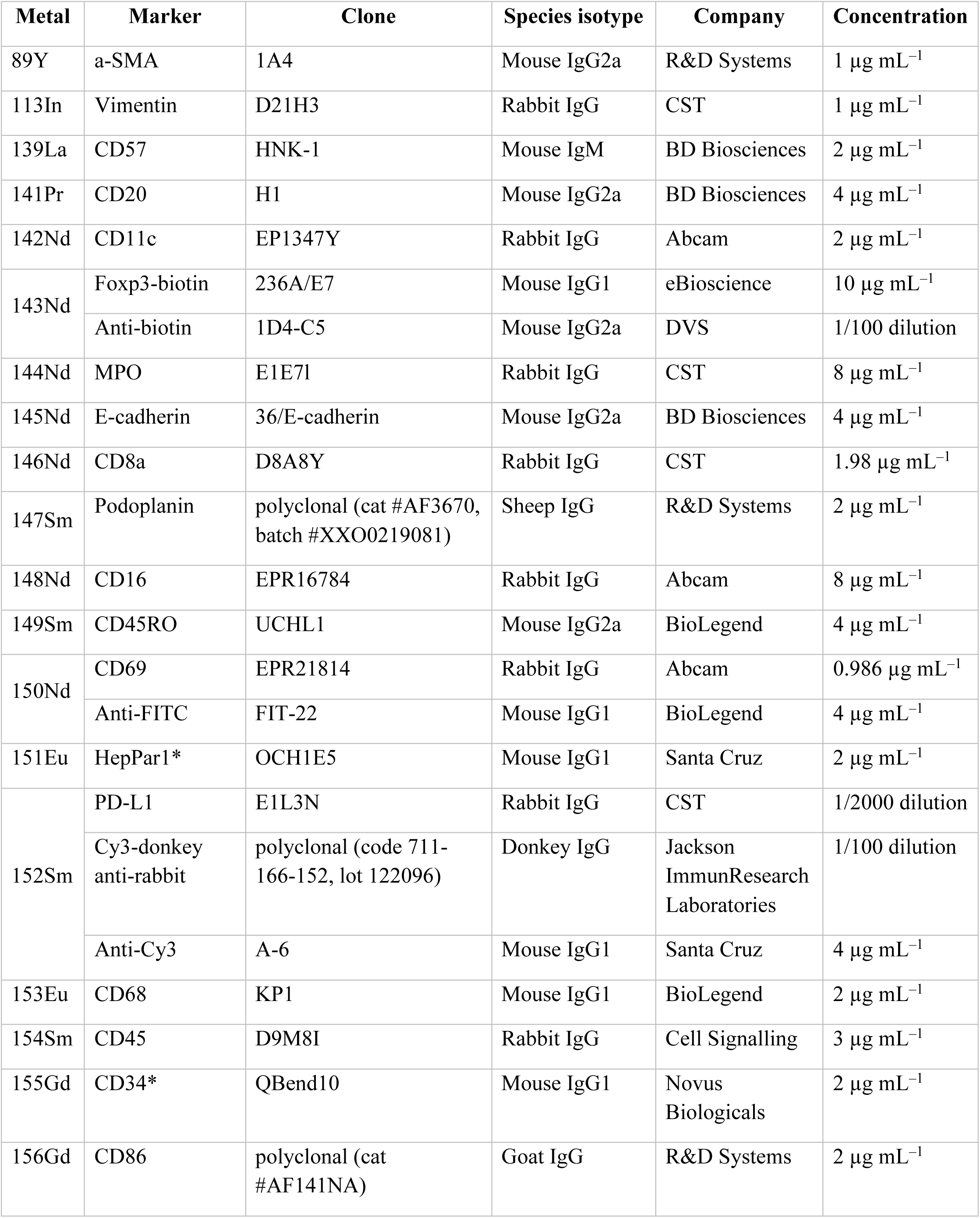
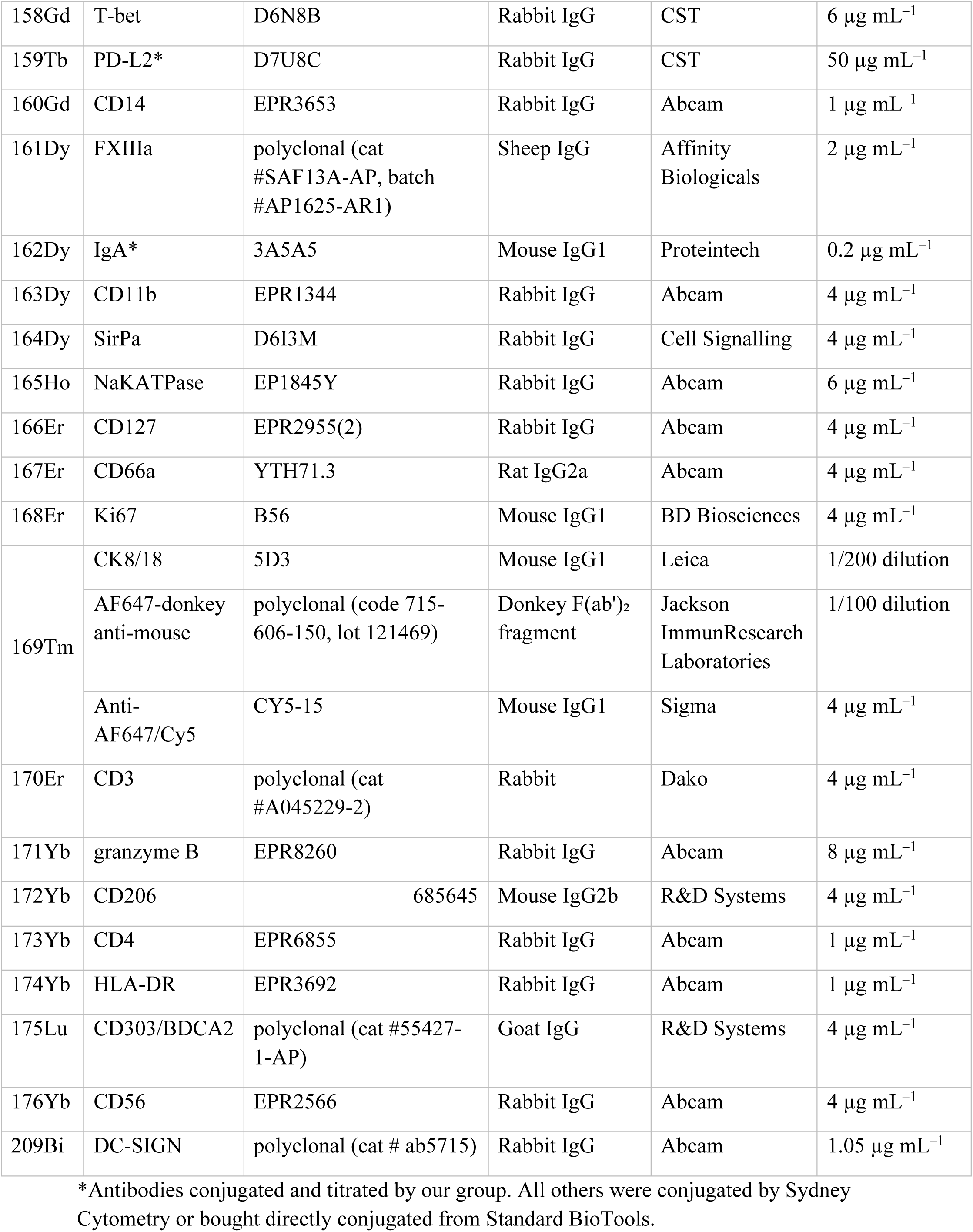
Imaging mass cytometry antibody panel.

